# Patient- and Family-Expressed Needs and Burdens in Kidney-Related Online Discussions in Taiwan: Large Language Model–Based Analysis Across Clinical Contexts

**DOI:** 10.64898/2026.11.17.26365963

**Authors:** Zhi-Yong Liu, Yu-Chi Hung, Chang-Fu Kuo

## Abstract

**Introduction:** What people with kidney concerns or disease and their families struggle with between visits may not surface in clinic. We mapped the needs and burdens they express online by clinical context and author.

**Methods:** A large language model read all 22,170 posts reached by a chronic kidney disease tag or kidney lexicon on a Taiwanese forum and 3 ask-a-physician services (1995–2026), applying a companion friction framework with kidney-specific fields, author (patient or family) and clinical context. A second vendor re-read 11,010 posts; 2 physicians labelled relevance, author and context in 200.

**Results:** Among 6,023 kidney-related posts, the reproducibly measured fields most often voiced on the forum were information needs (79%), emotional burden (48%), system navigation (34%) and worry about progression (24%). After abnormal tests, 54%–56% of posts quoted a laboratory value, nearly always with a question or stated uncertainty; renal-diet burden peaked with diagnosed disease; caregiver burden rose to 29% of dialysis-or-transplantation forum posts. Family-written posts more often voiced navigation difficulty and caregiver burden. Most fields agreed with the second vendor (κ 0.65–0.98). On the model’s labels, family members wrote 25% of forum posts about kidney concerns and 74% about dialysis or transplantation; on a 139-post sample, physicians’ labels gave 47%–48% for dialysis or transplantation (model 71%).

**Conclusion:** Kidney-related posts carry context-specific information needs and emotional and practical burdens, many voiced by relatives. They suggest candidate responses (test-result explanation, diet support, family-directed information and support) that need evaluation.

## Introduction

Chronic kidney disease (CKD) affects about one in ten adults worldwide,^1^ and Taiwan has among the highest incidence and prevalence of treated kidney failure.^2^ Much of its lived burden falls between visits and on families: caregivers of adults on dialysis report substantial burden and impaired quality of life,^3^ core outcome sets for hemodialysis trials include outcomes that patients and caregivers prioritize,^4^ and Kidney Disease: Improving Global Outcomes (KDIGO) places shared decision-making at the center of CKD care.^5^ Patient-reported outcome instruments capture predefined domains at scheduled times, but not what patients and families spontaneously struggle with: what a result means, what to eat, whether function will decline, how to arrange dialysis.

Public online posts offer an unprompted record of these difficulties, and guidance treats social-media data as a source of patient experience.^6^ Listening studies have described information needs and experiences in IgA nephropathy, CKD and dialysis in English, Korean and Chinese communities,^7–11^ and relatives search for health information on behalf of others;^12–16^ a kidney caregiver forum study described caregivers’ knowledge needs and burden.^17^ These studies described themes, but not how needs and burdens differ between a first abnormal test, diagnosed disease and dialysis, or between what patients and relatives write. A companion study defined an eight-domain framework of patient-expressed needs, burdens and barriers (“frictions”) for Taiwanese online discussions.^18^ Venue also matters: on ask-a-physician services every post is a question, whereas forums elicit narrative.^19^

We applied that framework, with kidney-specific fields, to every post that a CKD tag or a kidney lexicon reached on four Taiwanese platforms. We asked which needs and burdens kidney-related posts voice in each clinical context, whose they are (the patient’s or a relative’s), and who writes about each context.

## Methods

### Design and Timeline

This cross-sectional content analysis of public posts proceeded in a documented order (Supplementary Methods S1; Supplementary Tables S1–S3). The kidney frame was read first; rubrics for six comparison conditions were then frozen and hashed (SHA-256) before their posts were read, and an analysis plan centred on authorship was frozen (2026-10-25T08:46Z) after the context-by-author results had been inspected. The kidney friction profile was then made the primary result and the authorship comparison secondary; both had been inspected before this reprioritization, so all analyses are descriptive or exploratory, and the patient-versus-family and abnormal-test summaries were specified after the primary results had been inspected. Reporting follows the STROBE (Strengthening the Reporting of Observational Studies in Epidemiology) and TRIPOD-LLM (Transparent Reporting of a Multivariable Model for Individual Prognosis or Diagnosis–Large Language Models) guidelines^20,21^ (Supplementary Tables S10 and S11).

### Data Sources

Posts came from a research archive of four public Taiwanese platforms whose terms did not prohibit non-commercial academic analysis: PTT, an academically operated open forum,^22^ and three ask-a-physician services (KingNet, Taiwan e-Hospital of the Ministry of Health and Welfare, and tcm.tw), with hashed author identifiers. Platform terms and robots.txt were audited (16 August 2026). The companion study^18^ uses the same archive and ethics approval but a different sample and question and reports none of these results; of the 36,564 distinct posts read across the seven conditions (37,630 frame memberships), 338 were among its sampled units, 198 in the kidney frame. The unit was a forum article (one-line comments excluded) or a service submission, with appended physician replies, quoted text and signatures removed. Results are reported separately for the forum and the services.

### Kidney Frame and Reading

The kidney frame comprised every post that carried the archive pipeline’s CKD tag with its patient-voice flag (14,392 posts) or matched a 28-term Traditional Chinese kidney lexicon (13,650 posts; terms in Supplementary Methods S2): 22,170 distinct posts (forum 7,302; services 14,868), all of which were read. Posts using none of these terms were not reached, so the frame’s recall is unknown.

Every post was read in full by one large language model (gpt-6.1-sol, OpenAI, through its command-line interface; 10 posts per call; October 2026)^23,24^ under a relevance rubric with explicit rules: kidney disease counts when deleting it would change what the author asks or tells; stones, infection and kidney cancer count only when kidney function, dialysis or transplantation is at issue; and every rejection records a reason. A first read under an earlier rubric had rejected posts that physicians counted, so the rubric was revised and frozen before this read (Table 1; Supplementary Methods S3). The author was coded as patient, caregiver (a family member or friend writing about, or for, the patient; called family below, as friends wrote 4.4% of these posts), mixed, professional, media or other.

**Table 1.** The kidney frame, confirmation, and measurement checks by venue.

| Venue | Posts read | Confirmed kidney posts<sup>a</sup> | κ, original vs revised read | CKD-tag precision, % (95% CI)<sup>b</sup> | Analysed posts<sup>c</sup> | Family or friend author, % (95% CI) | Second-vendor re-read, n (%) | Second-vendor κ: relevance; author; context group |
|---|---|---|---|---|---|---|---|---|
| Forum (PTT articles) | 7,302 | 1,016 | 0.85 | 9.8 (8.7–10.9) | 1,013 | 53.3 (50.2–56.4) | 7,302 (100) | 0.85; 0.99; 0.92 |
| Ask-a-physician services<sup>d</sup> | 14,868 | 5,018 | 0.83 | 33.9 (33.0–34.8) | 5,010 | 29.7 (28.5–31.0) | 3,708 (25) | 0.90; 0.96; 0.90 |
| All | 22,170 | 6,034 | 0.84 | 28.9 (28.2–29.6) | 6,023 | 33.7 (32.5–34.9) | 11,010 (50) | — |

<sup>a</sup>Confirmed by the revised relevance read as concerning a specific person’s kidney disease or kidney concern and written by the patient, a family member or friend, or both (mixed). <sup>b</sup>Share of posts carrying the archive pipeline’s CKD tag and patient-voice flag that the revised read confirmed; on the three services separately, KingNet 30.8%, Taiwan e-Hospital 39.8% and tcm.tw 34.0%. <sup>c</sup>Confirmed posts written by the patient or by family or friends with a context assigned (mixed posts excluded). <sup>d</sup>KingNet, Taiwan e-Hospital and tcm.tw. κ, original vs revised read: agreement on confirmation between the first read and the revised, frozen read on all posts read. Second-vendor κ: Claude Sonnet 5.5 under the identical prompts; relevance on all re-read posts, author and context group on posts both readers confirmed. All shares are model estimates.
CKD, chronic kidney disease.

### Friction Framework and Kidney Fields

A content read applied the companion framework’s eight domains^18^ to confirmed posts: information gap (seeks information, asks a clinical question or expresses uncertainty), emotional burden, system navigation (insurance, paperwork, or which hospital or specialty to approach), access barrier, cost burden, caregiver burden (strain voiced by a helping relative), medication experience and lifestyle burden. Kidney-specific fields recorded renal-diet burden, worry about progression, dialysis concern and transplantation concern, and the relevance read recorded whether a kidney test value or stage was quoted. Each field is binary and assigned only when stated in the text.

Kidney context was read in six categories and grouped: concern (a kidney concern without a stated condition; an abnormal kidney test without a diagnosis), diagnosed (diagnosed CKD; another kidney condition with kidney function at issue) and dialysis or transplantation. These are cross-sectional posting contexts, not KDIGO stages or observed progression. Author and context were labelled in the same call, so cues to authorship may have influenced context. A descriptor read recorded whom the post concerned and whether it stated that the patient depended on others.

### Replication and Physician Labels

A second vendor (Claude Sonnet 5.5, Anthropic) re-read, under the identical frozen prompts, all kidney forum posts and a seeded quarter of kidney service posts (11,010 posts) and the content fields of all confirmed kidney posts. Two physician authors (YCH and CFK; CFK wrote the rubrics), blinded to model labels and to each other, personally read and labelled 200 kidney posts (no labels were pre-filled) drawn in purposive strata for relevance, author and context; these labels informed the rubric revision and are development material, not an independent validation, and they did not cover the friction fields.

### Outcomes and Statistical Analysis

The analysed population was posts confirmed as concerning a specific person’s kidney disease or concern and written by the patient or by family, with a context assigned; mixed posts were excluded. The primary result is the friction profile: the share of posts with each field, by context group and venue, with n/N and Wilson 95% CIs.^25^ Fields with second-vendor κ below 0.60 are shown but described as instrument-dependent. Within each context group, family-written and patient-written posts were compared as a Mantel–Haenszel risk difference across venues; these are different authors, not two views of the same patient. The secondary result is the family share of authorship by context group, with the dialysis-or-transplantation-minus-concern risk difference and Newcombe 95% CI,^26^ a 1-df logistic trend adjusted for board group and period (forum; author-clustered standard errors [SEs]) or site and period (services), and sensitivity analyses using the second vendor’s labels, posts both readers agreed on, mixed posts as family posts, the patient’s age group and the first post of each forum author. Six comparison conditions, read with the same template in seeded samples of up to 3,000 posts per condition (15,460 posts; posts per condition and venue in Supplementary Table S4), are reported in the Supplementary Material. Agreement is Cohen κ. Every number was generated by scripts from per-post label files.

### Ethics

The Institutional Review Board of Chang Gung Memorial Hospital approved the study (No. 202601393B0) and waived the requirement for informed consent. The study analysed lawfully public, non-interactive posts with hashed author identifiers; no one was contacted.^27,28^ No post text, searchable paraphrase or user identifier is reproduced. Post text, with author identifiers and any names, contact details or other identifiers within post bodies removed, was sent solely for labelling to OpenAI (Codex command-line interface) and Anthropic, under provider settings that exclude customer content from model training.

## Results

### Kidney-Related Posts

Of 22,170 posts read, 6,034 were confirmed as kidney-related patient or family voice; the commonest reasons for rejection were that the post was not about kidney disease (9,892), mentioned the kidney only in passing (2,566) or concerned no specific person (1,638) (Supplementary Table S4). After exclusion of 11 mixed posts, 6,023 were analysed: 1,013 forum articles by 846 authors and 5,010 service submissions (Table 1); 3,076 (51.1%) concerned a kidney concern or abnormal test, 1,899 diagnosed kidney disease (841 of them diagnosed CKD) and 1,048 dialysis or transplantation. Median publication years were 2014 (forum) and 2011 (services). The pipeline’s CKD tag was 9.8% precise on the forum (95% CI 8.7–10.9) and 30.8%–39.8% on the services, and the tagged frame reached 29% of confirmed forum posts. The second vendor agreed on relevance at κ 0.85 (forum) and 0.90 (services), on author at 0.99 and 0.96, on context group at 0.92 and 0.90, and on the friction and kidney fields at κ 0.65–0.98, except medication experience (0.52) and lifestyle burden (0.43).

### Needs and Burdens by Clinical Context

Kidney-related posts voiced a wide range of frictions (Table 2; Figure 1). Among reproducibly measured fields, on the forum 79% sought information or expressed uncertainty, 48% voiced emotional burden, 34% system navigation and 24% worry about progression; on the services, where every post is a question, 24% voiced emotional burden, 17% navigation and 28% worry about progression. Medication experience (36% and 38%) was common but instrument-dependent. Cost burden was labelled in at most 3.4% of posts in any context group and venue.

**Table 2.**
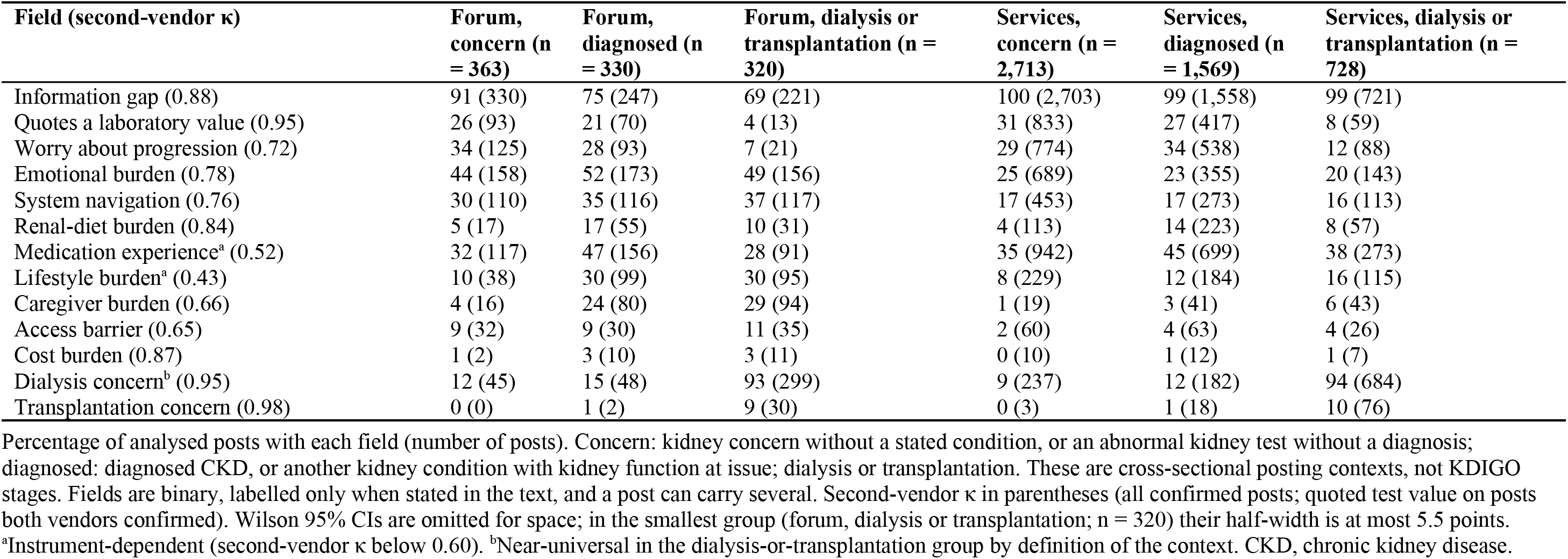
Needs and burdens voiced in kidney-related posts, by clinical context group and venue (model estimates).

**Figure 1.**
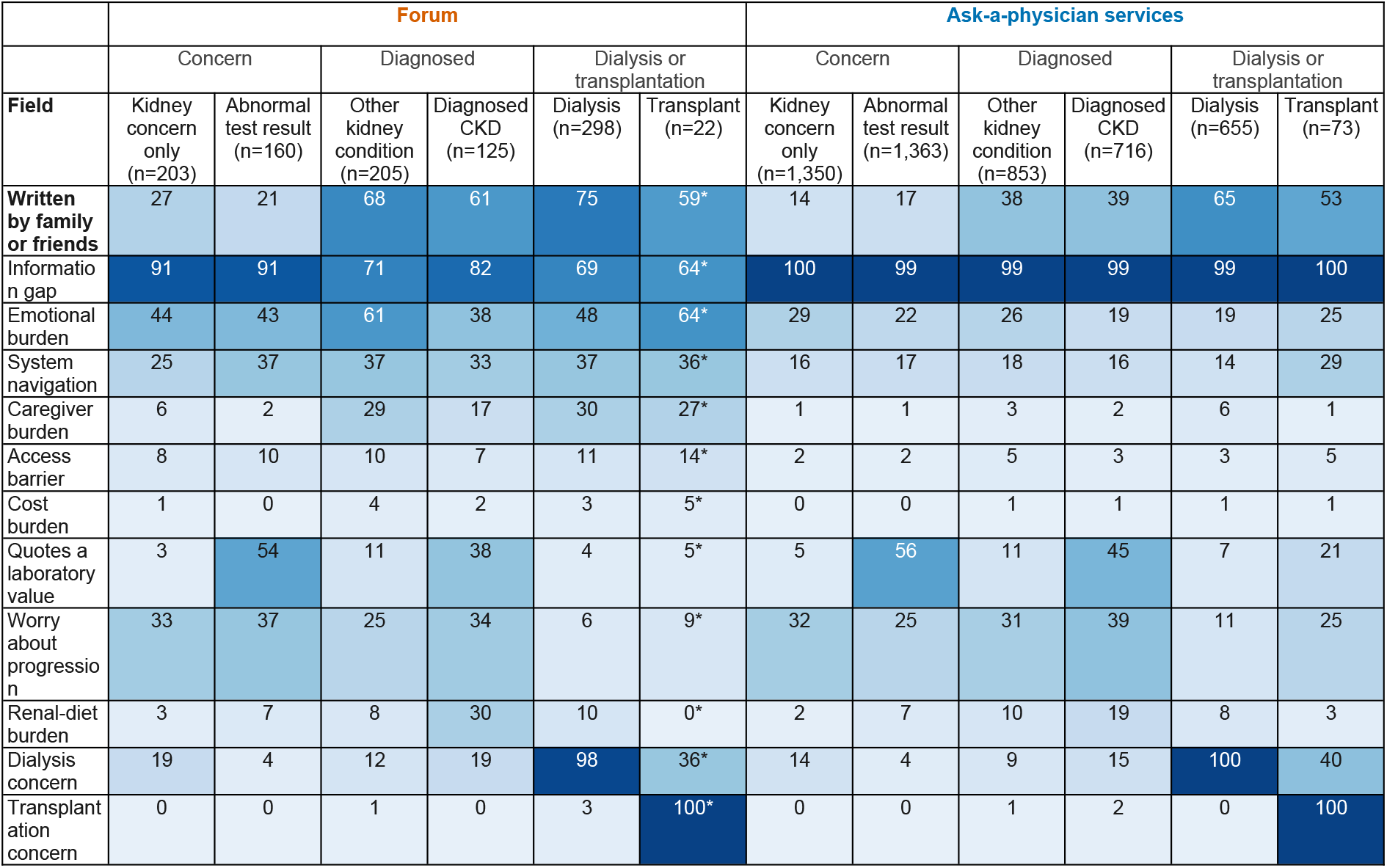
Needs and burdens voiced in kidney-related posts across six kidney contexts. Percentage of kidney-related posts (patient- and family-written) with each field, by context and venue; contexts are grouped into the concern, diagnosed and dialysis-or-transplantation groups. The top row is the share written by family members. Fields shown are those with second-vendor κ of at least 0.60; every field, with counts, is in Table 2. Asterisks mark contexts with fewer than 30 posts. All shares are model estimates.

The frictions differed by context. In the concern group, the dominant need was understanding a result: 91% of forum posts sought information, and 54% of forum and 56% of service posts about an abnormal test quoted a laboratory value, 94% and 99.6% of them together with a question or stated uncertainty; worry about progression was voiced in 34% and 29%. With diagnosed disease, daily management came forward: renal-diet burden rose to 17% (forum) and 14% (services) and was highest in posts about diagnosed CKD (30% and 19%; Figure 1), and emotional burden reached 52% of forum posts. In the dialysis-or-transplantation group, dialysis concern was near-universal by definition (93% and 94%); navigation (37% of forum and 16% of service posts) and emotional burden (49% and 20%) were as common as in other groups, and what distinguished the group was the relative’s own caregiver burden (29% and 6%), whereas quoting a test value and worry about progression became rare (4% and 7% on the forum). Medication experience (28%–47%) and lifestyle burden (8%–30%) were common in every group but instrument-dependent.

### Whose Needs and Burdens

Family-written posts voiced different frictions from patient-written posts in the same context group (Table 3). Family posts more often voiced system navigation in the concern (venue-adjusted difference +4.4 points, 95% CI +0.5 to +8.3) and dialysis-or-transplantation groups (+8.9, +4.0 to +13.7; diagnosed +2.1, −1.6 to +5.7), access barriers in the last group (+3.3, +0.7 to +5.9) and their own caregiver burden (+6.2 to +17.5 points). Patient-written posts more often quoted their own test values (diagnosed −8.1 points, 95% CI −12.2 to −4.1) and voiced lifestyle burden, an instrument-dependent field (diagnosed −3.5, −6.7 to −0.2; dialysis or transplantation −6.0, −11.2 to −0.7). On the forum, family posts about dialysis or transplantation more often sought information than patients’ posts (78% vs 45%) and voiced emotional burden (54% vs 33%), whereas patients more often voiced lifestyle burden (43% vs 25%). Renal-diet burden differed in opposite directions by venue: in the diagnosed group it was more common in family posts on the services (19% vs 11%) but in patients’ posts on the forum (22% vs 14%), so the pooled difference (+5.0 points) should not be read as a general family excess.

**Table 3.**
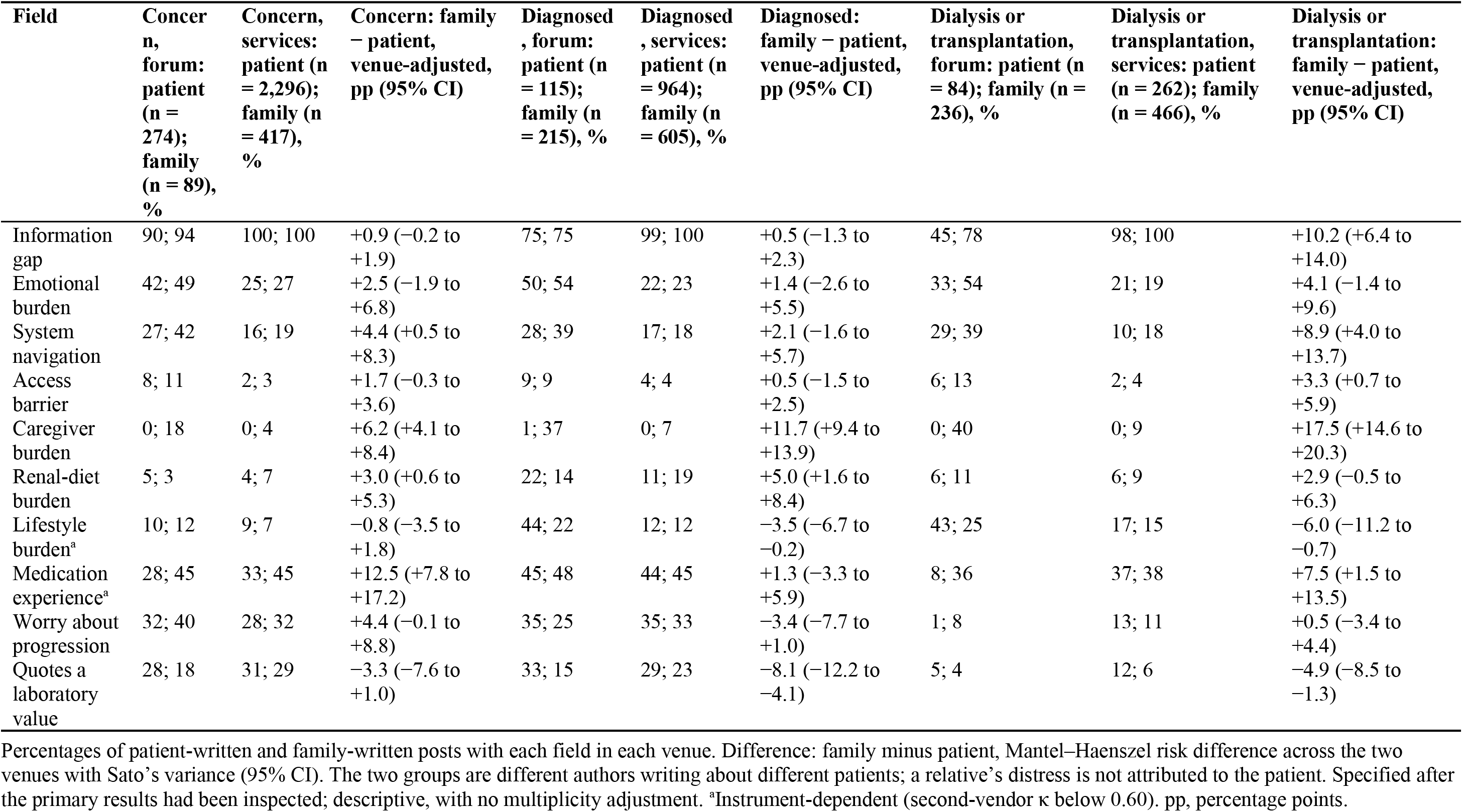
Needs and burdens in patient-written and family-written kidney-related posts within each context group (model estimates).

| **Field** | **Concern, forum: patient (n = 274); family (n = 89), %** | **Concern, services: patient (n = 2,296); family (n = 417), %** | **Concern: family − patient, venue-adjusted, pp (95% CI)** | **Diagnosed, forum: patient (n = 115); family (n = 215), %** | **Diagnosed, services: patient (n = 964); family (n = 605), %** | **Diagnosed: family − patient, venue-adjusted, pp (95% CI)** | **Dialysis or transplantation, forum: patient (n = 84); family (n = 236), %** | **Dialysis or transplantation, services: patient (n = 262); family (n = 466), %** | **Dialysis or transplantation: family − patient, venue-adjusted, pp (95% CI)** |
|---|---|---|---|---|---|---|---|---|---|
| Information gap | 90; 94 | 100; 100 | +0.9 (−0.2 to +1.9) | 75; 75 | 99; 100 | +0.5 (−1.3 to +2.3) | 45; 78 | 98; 100 | +10.2 (+6.4 to +14.0) |
| Emotional burden | 42; 49 | 25; 27 | +2.5 (−1.9 to +6.8) | 50; 54 | 22; 23 | +1.4 (−2.6 to +5.5) | 33; 54 | 21; 19 | +4.1 (−1.4 to +9.6) |
| System navigation | 27; 42 | 16; 19 | +4.4 (+0.5 to +8.3) | 28; 39 | 17; 18 | +2.1 (−1.6 to +5.7) | 29; 39 | 10; 18 | +8.9 (+4.0 to +13.7) |
| Access barrier | 8; 11 | 2; 3 | +1.7 (−0.3 to +3.6) | 9; 9 | 4; 4 | +0.5 (−1.5 to +2.5) | 6; 13 | 2; 4 | +3.3 (+0.7 to +5.9) |
| Caregiver burden | 0; 18 | 0; 4 | +6.2 (+4.1 to +8.4) | 1; 37 | 0; 7 | +11.7 (+9.4 to +13.9) | 0; 40 | 0; 9 | +17.5 (+14.6 to +20.3) |
| Renal-diet burden | 5; 3 | 4; 7 | +3.0 (+0.6 to +5.3) | 22; 14 | 11; 19 | +5.0 (+1.6 to +8.4) | 6; 11 | 6; 9 | +2.9 (−0.5 to +6.3) |
| Lifestyle burden<sup>a</sup> | 10; 12 | 9; 7 | −0.8 (−3.5 to +1.8) | 44; 22 | 12; 12 | −3.5 (−6.7 to −0.2) | 43; 25 | 17; 15 | −6.0 (−11.2 to −0.7) |
| Medication experience<sup>a</sup> | 28; 45 | 33; 45 | +12.5 (+7.8 to +17.2) | 45; 48 | 44; 45 | +1.3 (−3.3 to +5.9) | 8; 36 | 37; 38 | +7.5 (+1.5 to +13.5) |
| Worry about progression | 32; 40 | 28; 32 | +4.4 (−0.1 to +8.8) | 35; 25 | 35; 33 | −3.4 (−7.7 to +1.0) | 1; 8 | 13; 11 | +0.5 (−3.4 to +4.4) |
| Quotes a laboratory value | 28; 18 | 31; 29 | −3.3 (−7.6 to +1.0) | 33; 15 | 29; 23 | −8.1 (−12.2 to −4.1) | 5; 4 | 12; 6 | −4.9 (−8.5 to −1.3) |

Percentages of patient-written and family-written posts with each field in each venue. Difference: family minus patient, Mantel–Haenszel risk difference across the two venues with Sato’s variance (95% CI). The two groups are different authors writing about different patients; a relative’s distress is not attributed to the patient. Specified after the primary results had been inspected; descriptive, with no multiplicity adjustment. <sup>a</sup>Instrument-dependent (second-vendor κ below 0.60). pp, percentage points.

### Who Writes, by Clinical Context

On the model’s labels, family members wrote 53.3% of forum and 29.7% of service posts, with large differences between context groups (Table 4; Figure 2a): 24.5%, 65.2% and 73.8% of forum posts in the concern, diagnosed and dialysis-or-transplantation groups, and 15.4%, 38.6% and 64.0% of service posts. The dialysis-or-transplantation-minus-concern differences were +49.2 (95% CI 42.3 to 55.4) and +48.6 (44.8 to 52.3) percentage points (trend odds ratios 4.00 and 3.28 per group). The direction was reproduced with the second vendor’s own labels (+50.9 and +53.3 points), on posts both readers agreed on, with mixed posts counted as family posts, after adjustment for the patient’s age group, which was usually unstated (Supplementary Table S6), and with one post per forum author (26%, 67% and 82%) (Table 4; Supplementary Table S7). Family posts most often concerned a parent (49.0%, 54.1% and 59.8% across groups, venues pooled), with grandparents rising from 3.4% to 9.5%, whereas posts about the writer’s own child fell from 18.4% to 0.9% (Figure 2b); stated dependence of the patient rose from 7.5% to 15.0% and 27.4% (Figure 2c; trends in Supplementary Table S5). The one trend specified before its data were opened, a rising share of family posts saying that the patient was unable to ask, was not seen in kidney posts overall (pooled odds ratio per context group 1.04, 95% CI 0.91–1.18), although it was present on the forum (1.46, 1.13–1.90) and absent on the services (0.92, 0.79–1.07); in lung and colorectal cancer the pooled odds ratios were 3.00 and 2.93 (Supplementary Table S5).

**Table 4.** Family authorship of kidney-related posts by clinical context, with sensitivity analyses (model estimates unless stated).

| Context group and context | Forum, n | Forum, family or friend author, % (95% CI) | Services, n | Services, family or friend author, % (95% CI) |
|---|---|---|---|---|
| **Concern group** | **363** | **24.5 (20.4–29.2)** | **2,713** | **15.4 (14.1–16.8)** |
| Kidney concern without a stated condition | 203 | 27.1 (21.4–33.6) | 1,350 | 14.1 (12.3–16.0) |
| Abnormal kidney test result | 160 | 21.3 (15.6–28.2) | 1,363 | 16.7 (14.8–18.7) |
| **Diagnosed group** | **330** | **65.2 (59.9–70.1)** | **1,569** | **38.6 (36.2–41.0)** |
| Other kidney condition with kidney function at issue | 205 | 67.8 (61.1–73.8) | 853 | 38.2 (35.0–41.5) |
| Diagnosed CKD | 125 | 60.8 (52.0–68.9) | 716 | 39.0 (35.5–42.6) |
| **Dialysis or transplantation group** | **320** | **73.8 (68.7–78.3)** | **728** | **64.0 (60.5–67.4)** |
| Dialysis | 298 | 74.8 (69.6–79.4) | 655 | 65.2 (61.5–68.7) |
| Kidney transplantation | 22 | 59.1 (38.7–76.7) | 73 | 53.4 (42.1–64.4) |
| **Dialysis or transplantation minus concern, percentage points (95% CI)** |  | +49.2 (+42.3 to +55.4) |  | +48.6 (+44.8 to +52.3) |
| **Secondary: odds ratio per group (95% CI); Holm-adjusted P<sup>a</sup>** |  | 4.00 (3.08–5.19); <.001 |  | 3.28 (2.64–4.07); <.001 |
| *Sensitivity analyses: dialysis-or-transplantation minus concern (95% CI); odds ratio per group* |  |  |  |  |
| Second vendor's own relevance, author and context labels | 899 | +50.9 (+43.9 to +57.0); 5.02 | 1,238 | +53.3 (+45.6 to +60.1); 3.59 |
| Posts on which both readers agreed on all three fields | 783 | +52.6 (+45.1 to +59.1); 5.43 | 1,087 | +53.1 (+44.8 to +60.5); 3.70 |
| Mixed patient-and-family posts counted as family | 1,016 | +49.5 (+42.6 to +55.6); 4.04 | 5,018 | +48.6 (+44.7 to +52.2); 3.27 |
| Adjusted for the patient's age group (odds ratio per group) |  | 3.70 (2.81–4.87) |  | 3.72 (3.06–4.53) |
| First post of each forum author: concern / diagnosed / advanced, % | 846 | 26 / 67 / 82 |  | — |
| Physician A's own labels, 139 posts (venues pooled): concern / diagnosed / advanced, % (n/N; 95% CI); P for trend |  | 24 (8/33; 13–41) / 22 (8/36; 12–38) / 47 (22/47; 33–61); .02 |  |  |
| Physician B's own labels, 139 posts (venues pooled): concern / diagnosed / advanced, % (n/N; 95% CI); P for trend |  | 14 (4/28; 6–31) / 33 (13/40; 20–48) / 48 (19/40; 33–63); .004 |  |  |
| Model labels on posts from the same 139: concern / diagnosed / advanced, % (n/N) |  | 13 (4/31) / 43 (15/35) / 71 (17/24) |  |  |

Concern: kidney concern without a stated condition, or an abnormal kidney test without a diagnosis; diagnosed: diagnosed CKD, or another kidney condition with kidney function at issue; advanced: dialysis or kidney transplantation. These are cross-sectional posting contexts, not KDIGO stages. Shares with Wilson 95% CIs. Difference: advanced minus concern, Newcombe 95% CI. <sup>a</sup>Secondary summary: odds ratio per context group (scored 0, 1, 2) from logistic models adjusted for board group and period (forum; author-clustered SEs) or site and period (services; site-by-period clustered SEs; model-based SEs gave the same conclusions); Holm adjustment across the 8 gradient tests of the cross-condition analysis (Supplementary Table S9). Sensitivity rows give the advanced-minus-concern difference (95% CI) and the odds ratio per group, with the number of posts analysed. Physicians: each physician's own relevance, author and context labels on the 139 posts of the 200-post kidney sample from strata not selected on author or context (forum and services pooled; these labels informed the rubric), beside the model's labels on posts from the same sample; Cochran–Armitage trend test. CKD, chronic kidney disease.

**Figure 2.**
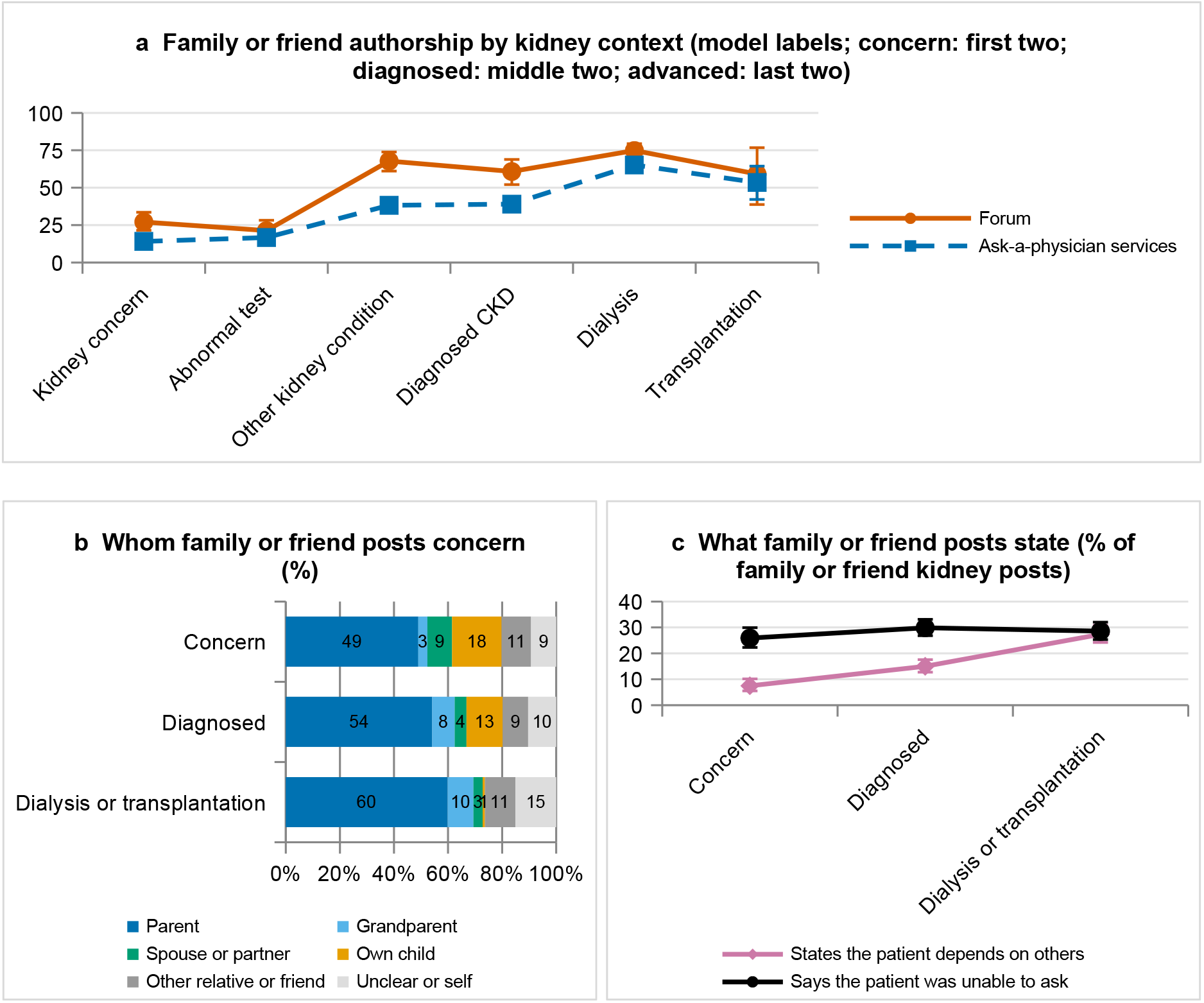
Who writes kidney-related posts, by clinical context. (a) Share of kidney-related posts written by family members in six kidney contexts, grouped into the concern, diagnosed and dialysis-or-transplantation context groups, on the forum (circles) and the ask-a-physician services (squares), with Wilson 95% CIs; open symbols mark contexts with fewer than 30 posts. Lines join cross-sectional groups and do not represent progression. All shares are model estimates. (b) Whom family posts concern, by context group (venues pooled). (c) Share of family posts stating that the patient depends on others, and saying that the patient was unable to ask, by context group (venues pooled; Wilson 95% CIs).

On the 139 physician-labelled posts not selected on author or context, each physician’s own labels gave a higher family share in the dialysis-or-transplantation than in the concern group, but smaller shares than the model in the diagnosed and dialysis-or-transplantation groups: physician A 24% (8/33), 22% (8/36) and 47% (22/47), *P* for trend=.02; physician B 14% (4/28), 33% (13/40) and 48% (19/40), *P*=.004; model labels on the same sample 13%, 43% and 71%. The direction is therefore better supported than the magnitude. Against the physicians, the author label agreed at κ 0.73 and 0.76 (between physicians 0.93) and the context group at 0.48 and 0.58 (0.69); physicians drew the boundary of a kidney post more widely than the model and agreed poorly on it (κ 0.18 and 0.38 with the model; 0.36 between physicians) (Supplementary Table S8). In exploratory comparisons, posts about advanced lung and colorectal cancer and interstitial lung disease were also mostly family-written, whereas posts about polycystic ovary syndrome and obesity stayed patient-written (3.1%–5.0%) (Supplementary Table S9; Supplementary Figure S1).

## Discussion

Reading every post that a kidney tag or lexicon reached, under an explicit framework, showed what people with kidney concerns or disease and their families struggle with online, and how this changes with clinical context. After an abnormal test the need was to understand a result, usually with the value quoted and a question asked, often with worry about progression. With diagnosed disease, renal-diet burden came forward, alongside medication and lifestyle burden, which were instrument-dependent. Around dialysis or transplantation, navigation and distress remained common and the distinctive friction was the strain of the relative who helps. Relatives wrote most of the posts about dialysis or transplantation on the model’s labels, and nearly half on the physicians’, and their posts more often voiced navigation difficulty (and, around dialysis, access difficulty), while patients’ own posts more often quoted their test values.

These are expressed concerns, not measured unmet needs, but they point to candidate responses that nephrology services can evaluate. Patient-readable explanation of kidney results, including what a value means and what to ask next, matches the dominant need after an abnormal test. Practical, context-appropriate dietary support matches the renal-diet burden of diagnosed disease. Information on services, appointments and where to get help matches the navigation voiced in every context, and around dialysis it should be written so that the relatives who ask can use it, with support for their own strain. KDIGO frames CKD care around shared decision-making,^5^ and Taiwan’s pre-end-stage kidney disease care program is designed around patients with advanced CKD;^29^ relatives’ burden in dialysis is substantial^3^ and their priorities enter core outcome sets.^4^ Providing information and support for relatives, while asking patients whom they want involved, is consistent with growing provision for care partners in patient portals;^30,31^ these posts do not show that patients cannot take part, and relatives should not replace patients in decisions. Cost burden was rarely labelled, as in the companion study,^18^ where physician agreement on cost was poor; universal coverage under National Health Insurance,^32^ selection of who posts and reporting practices may each contribute.

Previous nephrology listening studies^7^ described themes without separating context or author,^8–11^ and studies of relatives’ online writing in cancer^33^ and on a kidney caregiver forum^17^ sampled relatives’ posts directly. Our contribution is a context-resolved map of kidney frictions with the author identified: a listening study of dialysis posts that does not separate authors would largely describe relatives’ concerns, and a study defined by the pipeline’s CKD tag, correct for about one forum post in ten, would have described a different population.^18^

### Limitations

First, rates describe what was posted on four Taiwanese platforms, not people with kidney disease: half of the analysed posts concerned a kidney concern or abnormal test, context was inferred from the post, the frame’s recall is unknown, prolific authors may be over-represented, older adults are probably under-represented, and how often a burden is voiced is not its severity or evidence that care missed it. Second, all fields are model labels. Second-vendor agreement was high for most fields, but it shows reproducibility rather than validity; medication experience and lifestyle burden were instrument-dependent. The physicians labelled relevance, author and context but not the friction fields, agreed moderately on context and poorly on relevance, gave smaller family shares than the model, are authors, and informed the rubric revision, so they are not an independent check of the model’s labels. Third, the framework’s domains were defined in advance and frictions outside them were not measured; dialysis concern partly restates the context definition. Fourth, author and context were labelled in one pass, and the authorship and comparison-condition patterns were seen before the analyses were formalized, so all comparisons are exploratory. Fifth, posts span three decades (median years 2014 and 2011), the patient’s age group was usually unstated, and the data come from one health system and language. Finally, patients and caregivers did not help define the fields, and some posts may overlap with the models’ training data.

### Conclusion

Kidney-related posts on four Taiwanese platforms carry context-specific needs and burdens: understanding results after an abnormal test, renal diet with diagnosed disease, and family strain around dialysis or transplantation, with navigation common throughout and more often voiced by relatives. These concerns may not surface in routine visits; they identify candidate responses for patients and the relatives who help them, which require evaluation.

## Data Availability

All aggregate data, per-unit LLM label checkpoints, platform-terms audit snapshots, verbatim prompts, and analysis code will be deposited in a public repository (Zenodo) and are available upon reasonable request from the corresponding author. Raw post text is not redistributed due to platform terms of service and privacy protections, and the mapping from study identifiers to platform post identifiers is available to qualified researchers under a data-use agreement.

## Appendix

### List of Supplementary Material

Supplementary Methods S1. Study timeline, rubric hashes, analysis plan and deviations (Tables S1–S3).

Supplementary Methods S2. Kidney lexicon, context definitions and flow of posts (Table S4).

Supplementary Methods S3. Instrument history and frozen prompts.

Supplementary Results S1. Trends in family or friend posts and the patient’s age group (Tables S5–S6).

Supplementary Results S2. Sensitivity analyses and physician labels (Tables S7–S8) and the seven-condition comparison (Table S9; Supplementary Figure S1).

Supplementary Checklists. STROBE and TRIPOD-LLM (Tables S10–S11).

## Acknowledgments

We thank the patient and family communities whose public posts made this research possible. This work received no specific funding. Contributor roles (CRediT): CFK, conceptualization, methodology, formal analysis, investigation (physician labels), data curation, visualization, writing – original draft, writing – review and editing, supervision; YCH, investigation (physician labels), writing – review and editing; ZYL, data curation, resources, writing – review and editing. CFK had full access to all data and takes responsibility for the integrity of the data and the accuracy of the analysis.

## Declaration of Generative AI and AI-Assisted Technologies

Large language models served as study instruments (gpt-6.1-sol for the primary reads; Claude Sonnet 5.5 for the second-vendor reads), as described in the Methods; the frozen prompts are in the Supplementary Material. During manuscript preparation the authors used Anthropic Claude (Claude Code and Cowork, Claude Opus models) to assist with analysis code, drafting and editing; the authors reviewed and verified the code and every number and statement, and take full responsibility for the content.

## Data Sharing Statement

The data supporting this study are available from the corresponding author on reasonable request. Post text is not shared, because the platforms’ terms prohibit redistribution of user content.

## Supplementary Material

No post text, paraphrase identifiable by search or user identifier is reproduced in this material. Tables are numbered continuously (Tables S1–S11); Supplementary Figure S1 shows the seven-condition comparison. In the prompts, ‘stage’ corresponds to the context group of the main text and ‘caregiver’ to a family or friend author.

### Supplementary Methods S1. Study timeline, rubric hashes, analysis plan and deviations

**Table S1.** Order of work.

| Step | What was done | What had been seen at that point |
|---|---|---|
| 1 | Kidney frame (22,170 posts) read under the original full-text rubric; physician check (200 posts) | Kidney relevance, author and context labels |
| 2 | Kidney relevance rubric revised and frozen; kidney frame re-read; second-vendor re-read; kidney content read and its second-vendor re-read (descriptive) | Kidney family-or-friend share rising with kidney context (the context-group gradient) |
| 3 | Six-condition relevance, construct and descriptor rubrics written from the revised kidney rubric and frozen with SHA-256 hashes (2026-10-25T04:08Z) | No six-condition output |
| 4 | Six-condition relevance/author/context-group read (15,460 posts) and second-vendor re-read (40% of documents) | Context-group-by-author shares of all seven conditions (inspected) |
| 5 | Analysis plan version 1 written and frozen (2026-10-25T08:46:22Z) | As step 4; descriptor and construct outputs not opened |
| 6 | Descriptor read (all confirmed posts of seven conditions) and construct read (six conditions) opened and analysed under the plan | — |
| 7 | Kidney and comparison-condition analyses reported together in this paper; deviations recorded in Table S3 | — |

Consequence for interpretation: H1 (gradient) and H2 (equivalence) formalize, with a stated test, multiplicity correction and margin, a pattern that had been observed at step 4; they are not independent confirmatory tests. The H3 predictions (age group as mediator; rising share of ‘unable to ask’ within family-or-friend posts) were fixed at step 5, before their data were opened.

**Table S2.** Rubrics, prompts and scripts frozen before reading.

| **Rubric, prompt or script** | **Frozen (UTC)** |
|---|---|
| **Revised kidney rubrics**: |  |
| Revised kidney relevance, author and context rubric | 2026-10-24 15:35 |
| Revised kidney relevance prompt as sent | 2026-10-24 15:35 |
| Kidney construct prompt as sent | 2026-10-24 15:35 |
| Original kidney read prompt as sent | 2026-10-24 15:35 |
| Kidney re-read script | 2026-10-24 15:35 |
| **Six further conditions and descriptor read**: |  |
| **Rubric, prompt or script** | **Frozen (UTC)** |
| Descriptor rubric | 2026-10-25 04:08 |
| Condition definitions (six conditions) | 2026-10-25 04:08 |
| Relevance, author and stage template (six conditions) | 2026-10-25 04:08 |
| General construct rubric (six conditions) | 2026-10-25 04:08 |
| Relevance prompt as sent: vertigo | 2026-10-25 04:08 |
| Relevance prompt as sent: obesity | 2026-10-25 04:08 |
| Relevance prompt as sent: polycystic ovary syndrome | 2026-10-25 04:08 |
| Relevance prompt as sent: lung cancer | 2026-10-25 04:08 |
| Relevance prompt as sent: colorectal cancer | 2026-10-25 04:08 |
| Relevance prompt as sent: interstitial lung disease | 2026-10-25 04:08 |
| Construct prompt as sent (six conditions) | 2026-10-25 04:08 |
| Descriptor prompt as sent | 2026-10-25 04:08 |
| Six-condition read script | 2026-10-25 04:08 |

#### Analysis plan

The analysis plan (version 1; frozen 2026-10-25T08:46:22Z, before the descriptor and six-condition construct outputs were opened) and the SHA-256 hashes of the plan and of the rubrics, prompts and scripts in Table S2 are available with the data on request from the corresponding author. In the plan, H1 is the gradient comparison, H2 the comparison with the ±10-point margin and H3 the descriptor predictions; H1 and H2 formalize a pattern inspected before the plan was frozen and are exploratory, and only H3 was specified blind. H2 was reported descriptively rather than with its planned equivalence test: the 95% CIs of the diagnosed-minus-concern differences lay within the ±10-point margin in three of the four venue cells of the two usually self-managed conditions, and the lower limit for polycystic ovary syndrome (PCOS) on the forum was −10.1 points (Table S9).

**Table S3.** Deviations from the plan.

| # | Plan | What was done | Effect |
|---|---|---|---|
| 1 | CKD gradient to be reported in a separate kidney paper and shown here for comparison only | The kidney and comparison-condition analyses are reported together in this paper; the CKD gradient is reported here, identified as observed before the plan | None on estimates; CKD remains in the 8-test Holm family as planned |
| 2 | 1-df logistic trend test in every H1 cell | Forum interstitial lung disease (ILD) showed complete separation; a Fisher exact test of the merged contrast replaced the trend test. A coding error initially applied the exact test to services ILD as well; this was corrected so that services ILD uses the planned logistic test | All 8 Holm-adjusted P<.001 under either version |
| 3 | H3(ii) within-family-or-friend trend pooled over venues and stratified by venue | Pooled (venue-adjusted) trends in the main text; venue-stratified trends added in Supplementary Results S1 (Table S5) | Reported in full |
| 4 | Display items: Figure 2 risk differences with H2 margin; Figure 3 mechanism (plan numbering) | Risk differences in Table 4 and Supplementary Table S9; Figure 1 shows kidney content; Figure 2 who writes and kidney descriptors; Supplementary Figure S1 the seven conditions | Presentation only |
| 5 | Physician check of about 410 posts with adjudication | Not done; physician labels are available only for the 200 kidney posts (Table S8) | Context-group validity outside kidney disease rests on the second-vendor sensitivity analysis (Table S7) only |
| 6 | Kidney cohort of 6,034 posts; descriptor read of 6,037 posts | Analysed kidney population 6,023 (6,034 confirmed minus 11 mixed); the descriptor read covers all 6,023 | Single kidney denominator throughout |
| 7 | Any context-group-by-venue cell with fewer than 20 posts merged with the adjacent lower group | Only advanced cells were merged; the forum ILD concern cell (15 posts), which has no lower group, was retained as the H1 reference cell | Disclosed in Supplementary Table S9 and the Supplementary Figure S1 legend |

### Supplementary Methods S2. Kidney lexicon, context definitions and flow of posts

**Condition-specific definitions (English text of the frozen fill-ins of the rubric template; Chinese terms omitted)**

#### Lung cancer

*Counts:* (a) this person’s lung cancer, diagnosed or suspected, at any stage, including treatment, follow-up, recurrence, palliative and end-of-life care; (b) a lung nodule, ground-glass opacity or lung shadow under evaluation for possible cancer, and an abnormal lung-cancer screening result for this person; (c) questions about this person’s lung-cancer tests, treatment, side-effects, costs or prognosis. *Excluded:* cancer elsewhere that has spread to the lung when the post is about the other cancer, lung infection or other lung disease with no cancer concern. *Concern:* a nodule, shadow or screening finding under evaluation, or other symptoms, a test or imaging finding, or a question whether a drug, supplement or food harms this person, without a clinician-confirmed diagnosis. *Diagnosed:* confirmed lung cancer under treatment or follow-up that is not described as advanced. *Advanced:* stage IV or metastatic disease, recurrence, or treatment described as palliative (non-curative) intent.

#### Colorectal cancer

*Counts:* (a) this person’s colorectal cancer, diagnosed or suspected, at any stage, including surgery, a stoma, chemotherapy, follow-up, recurrence, palliative and end-of-life care; (b) blood in the stool, a positive faecal occult blood test, polyps or colonoscopy findings when colorectal cancer is a concern for this person; (c) questions about this person’s colorectal-cancer screening, tests, treatment, side-effects, costs or prognosis. *Excluded:* haemorrhoids, irritable bowel or other bowel complaints with no cancer concern, cancer elsewhere when the post is about the other cancer, a routine negative colonoscopy with no cancer concern. *Concern:* a positive screening test, polyps or a colonoscopy finding, or other symptoms, a test or imaging finding, or a question whether a drug, supplement or food harms this person, without a clinician-confirmed diagnosis. *Diagnosed:* confirmed colorectal cancer under treatment or follow-up that is not described as advanced. *Advanced:* stage IV or metastatic disease, recurrence, or treatment described as palliative (non-curative) intent.

#### Interstitial lung disease

*Counts:* (a) this person’s interstitial lung disease or pulmonary fibrosis, diagnosed or suspected, including idiopathic pulmonary fibrosis, interstitial pneumonia linked to autoimmune disease, occupation, drugs or COVID-19; (b) a CT or X-ray report mentioning fibrosis or interstitial change for this person that the author asks or worries about; (c) questions about this person’s ILD tests, antifibrotic or other treatment, oxygen, transplant evaluation or prognosis. *Excluded:* liver fibrosis, cystic fibrosis, lung cancer or infection without fibrosis or interstitial disease, a single mention of scarring or old fibrosis described by a clinician as insignificant with no follow-up requested. *Concern:* an imaging finding, or other symptoms, a test or imaging finding, or a question whether a drug, supplement or food harms this person, without a clinician-confirmed diagnosis. *Diagnosed:* ILD or pulmonary fibrosis diagnosed by a clinician or under treatment, not described as advanced. *Advanced:* prescribed home oxygen, antifibrotic therapy with stated progression, or lung-transplant evaluation.

#### Vertigo

*Counts:* (a) dizziness, vertigo, spinning, light-headedness or imbalance that this person has, has had repeatedly, or is worried about, including BPPV, Ménière’s disease, vestibular neuritis, vestibular migraine and unexplained or positional dizziness; (b) tests, examinations, medicines or treatment for this person’s dizziness (vestibular tests, hearing tests, imaging, repositioning manoeuvres, anti-vertigo drugs); (c) dizziness as the presenting complaint being evaluated when a cause such as low blood pressure, anaemia, heart rhythm or anxiety is being considered. *Excluded:* dizziness only as a metaphor, motion sickness, a hangover, a drug-leaflet side-effect list, or a one-word symptom in a list when the post is about something else; posts whose subject is an already-diagnosed stroke, dementia or other neurological illness in which dizziness is one symptom among others. *Concern:* dizziness or vertigo symptoms, a test or imaging finding, or a question whether a drug, supplement or food harms this person, without a clinician-confirmed diagnosis. *Diagnosed:* a named cause has been given by a clinician (BPPV, Ménière’s disease, vestibular neuritis, vestibular migraine, or another named diagnosis) or the dizziness is under ongoing treatment prescribed by a clinician. *Advanced:* a vestibular or central cause of the dizziness confirmed by a specialist that requires procedural treatment (inner-ear injection or surgery), or a specialist diagnosis of a central cause of the dizziness itself.

#### Polycystic ovary syndrome

*Counts:* (a) this person’s polycystic ovary syndrome, diagnosed or suspected, including polycystic ovaries on ultrasound; (b) irregular or absent periods, anovulation, hirsutism, acne or weight gain when PCOS is named or asked about as the cause; (c) PCOS treatment, fertility treatment or pregnancy planning in PCOS, and PCOS-related metabolic questions (insulin resistance, weight) for this person. *Excluded:* polycystic kidney disease or polycystic liver disease, other ovarian cysts (chocolate cysts, dermoid cysts) without PCOS, irregular periods with no mention of PCOS. *Concern:* suspected PCOS: symptoms, a test or imaging finding, or a question whether a drug, supplement or food harms this person, without a clinician-confirmed diagnosis. *Diagnosed:* PCOS diagnosed by a clinician or under treatment. *Advanced:* assisted reproduction (ovulation induction, IUI or IVF) under way or scheduled for this person because of PCOS, or endometrial hyperplasia or type 2 diabetes attributed to PCOS under treatment.

#### Obesity

*Counts:* (a) this person’s overweight or obesity, stated, believed or diagnosed, as a health concern (stated health reasons, not appearance alone); (b) this person’s efforts to lose weight for health reasons, including weight-loss medicines (including GLP-1 injections), a weight-loss clinic, dietitian care, or bariatric surgery; (c) obesity-related health problems (fatty liver, diabetes, sleep apnoea, joint pain) when this person’s weight is part of what is asked or described, in adults or children. *Excluded:* pregnancy or infant weight, muscle gain or fitness training without a weight problem, eating disorders and wanting to gain weight, cosmetic body shaping without a weight problem, appearance-only slimming goals. *Concern:* self-described overweight, or a test or imaging finding, or a question whether a drug, supplement or food harms this person, without a clinician-confirmed diagnosis, without medical treatment for weight or a stated BMI of 27 or more. *Diagnosed:* obesity stated by a clinician, a stated BMI of 27 or more, or medical treatment for weight (prescribed medicine, a weight-loss clinic, dietitian care). *Advanced:* bariatric surgery (done, scheduled or being evaluated), or a stated obesity-related complication for which this person receives prescribed treatment and which the author attributes to the weight (for example sleep apnoea on a breathing machine because of the weight).

#### Chronic kidney disease

Six contexts read by the revised kidney rubric and grouped into context groups: kidney concern without a stated condition and abnormal kidney test result (concern); diagnosed CKD and another kidney condition with kidney function at issue (diagnosed); dialysis and kidney transplantation (advanced). Stones, infection and kidney cancer count only when kidney function, kidney damage, dialysis or transplantation is at issue.

#### Kidney lexicon and frame composition

The kidney frame was the union of posts carrying the archive pipeline’s CKD tag with its patient-voice flag (14,392 posts) and posts matching a 28-term kidney lexicon in title or body (13,650 candidates on 2026-09-29), 22,170 distinct posts. Terms: disease names 腎臟病 (kidney disease), 慢性腎 (chronic kidney), 腎病變 (nephropathy), 腎功能 (kidney function), 腎衰竭 (kidney failure), 腎絲球 (glomerulus), 腎臟功能 (kidney function, long form); dialysis 洗腎 (dialysis, colloquial), 透析 (dialysis), 瘻管 (fistula), 廔管 (fistula, variant character); transplantation 換腎 (kidney transplant, colloquial), 腎臟移植 (kidney transplantation), 腎移植 (kidney transplant); laboratory 肌酸酐 (creatinine), 蛋白尿 (proteinuria), eGFR, GFR, 尿毒 (uraemia), 微白蛋白 (microalbumin), 泡泡尿 (foamy urine); other 腎臟科 (nephrology department), 腎病 (kidney disease, short form), 腎炎 (nephritis), 腎萎縮 (kidney atrophy), 腎不好 (bad kidneys), 腎臟不好 (kidneys not good), 顧腎 (protecting the kidneys). Of the confirmed kidney posts, the tagged frame reached 29% on the forum and 77% on the services; posts that used neither the tag vocabulary nor the lexicon were not reached, so recall of all kidney-related posts is unknown.

**Table S4.**
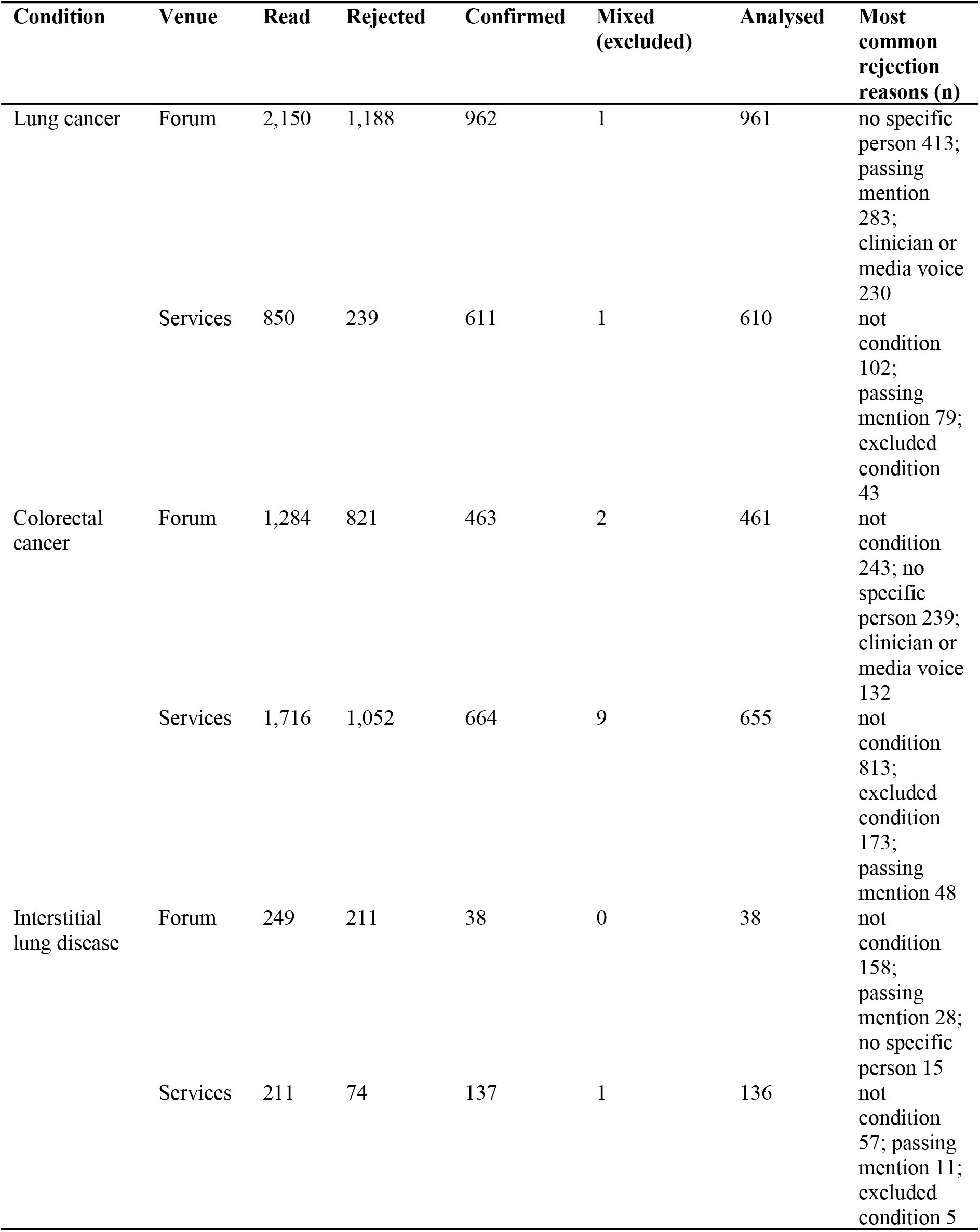
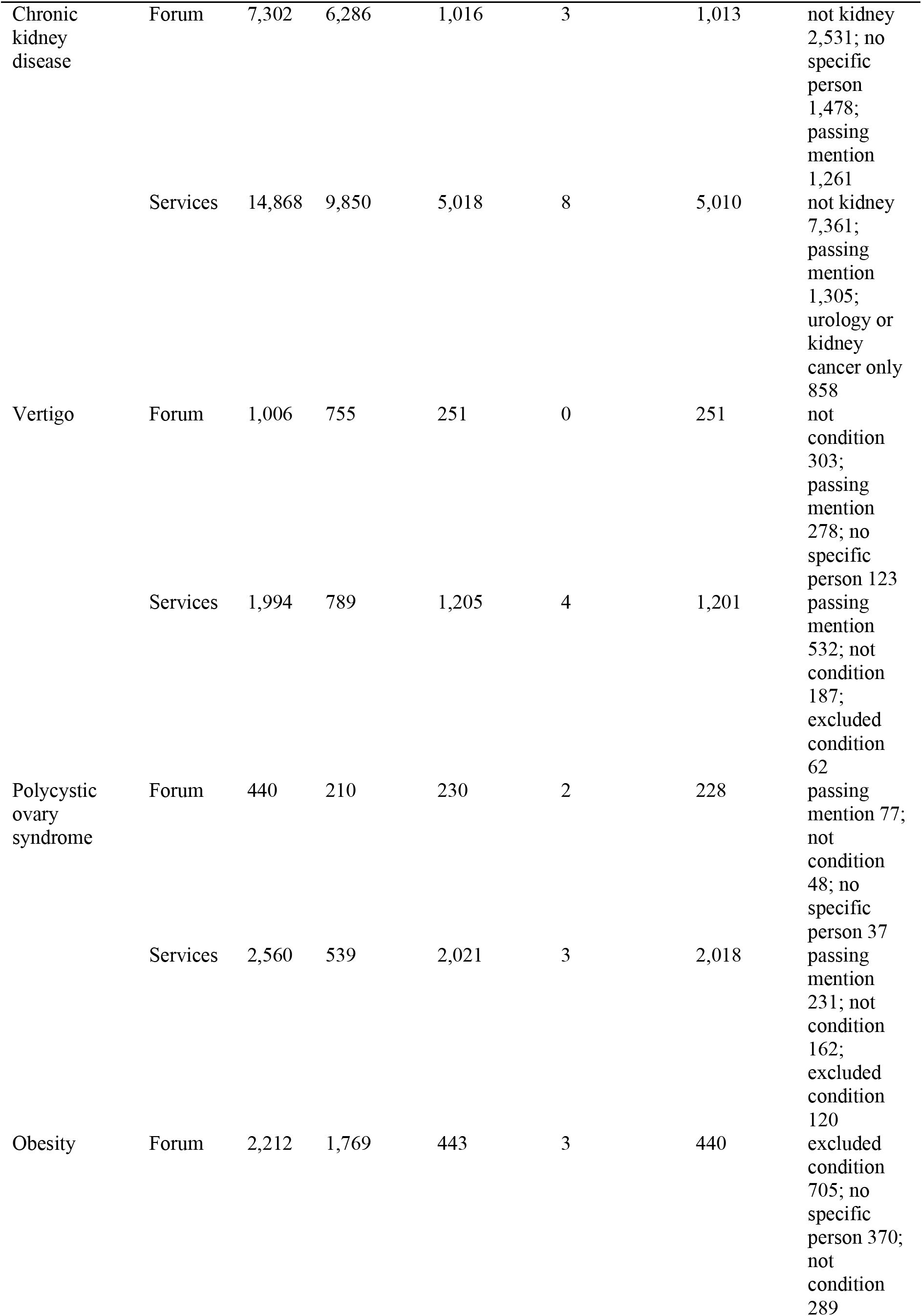
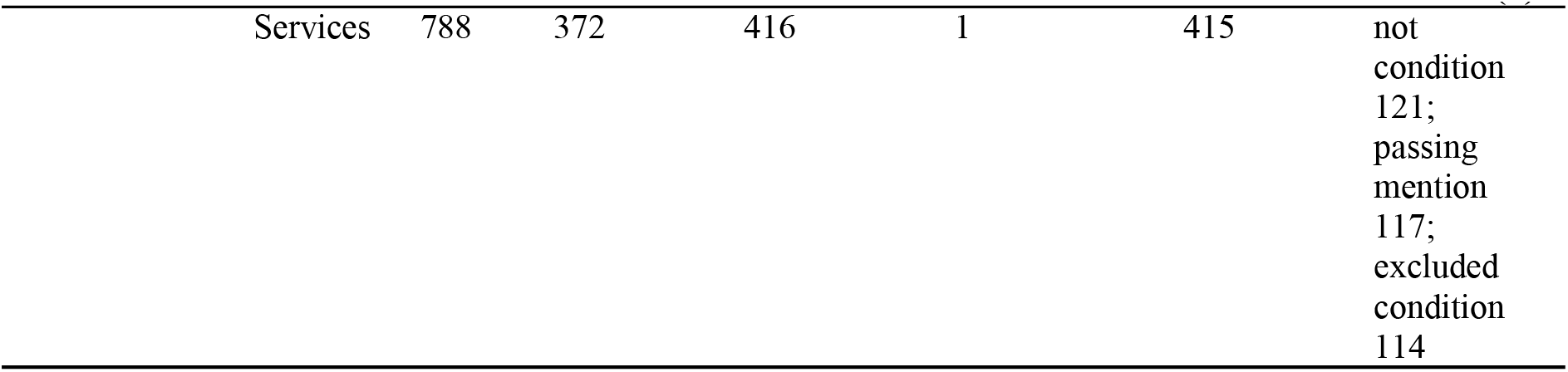
Flow of posts.

| **Condition** | **Venue** | **Read** | **Rejected** | **Confirmed** | **Mixed (excluded)** | **Analysed** | **Most common rejection reasons (n)** |
|---|---|---|---|---|---|---|---|
| Lung cancer | Forum | 2,150 | 1,188 | 962 | 1 | 961 | no specific person 413; passing mention 283; clinician or media voice 230 |
|  | Services | 850 | 239 | 611 | 1 | 610 | not condition 102; passing mention 79; excluded condition 43 |
| Colorectal cancer | Forum | 1,284 | 821 | 463 | 2 | 461 | not condition 243; no specific person 239; clinician or media voice 132 |
|  | Services | 1,716 | 1,052 | 664 | 9 | 655 | not condition 813; excluded condition 173; passing mention 48 |
| Interstitial lung disease | Forum | 249 | 211 | 38 | 0 | 38 | not condition 158; passing mention 28; no specific person 15 |
|  | Services | 211 | 74 | 137 | 1 | 136 | not condition 57; passing mention 11; excluded condition 5 |
| Chronic kidney disease | Forum | 7,302 | 6,286 | 1,016 | 3 | 1,013 | not kidney 2,531; no specific person 1,478; passing mention 1,261 |
|  | Services | 14,868 | 9,850 | 5,018 | 8 | 5,010 | not kidney 7,361; passing mention 1,305; urology or kidney cancer only 858 |
| Vertigo | Forum | 1,006 | 755 | 251 | 0 | 251 | not condition 303; passing mention 278; no specific person 123 |
|  | Services | 1,994 | 789 | 1,205 | 4 | 1,201 | passing mention 532; not condition 187; excluded condition 62 |
| Polycystic ovary syndrome | Forum | 440 | 210 | 230 | 2 | 228 | passing mention 77; not condition 48; no specific person 37 |
|  | Services | 2,560 | 539 | 2,021 | 3 | 2,018 | passing mention 231; not condition 162; excluded condition 120 |
| Obesity | Forum | 2,212 | 1,769 | 443 | 3 | 440 | excluded condition 705; no specific person 370; not condition 289 |
|  | Services | 788 | 372 | 416 | 1 | 415 | not condition 121; passing mention 117; excluded condition 114 |

#### Kidney cohort reconciliation

The revised kidney read confirmed 6,034 posts as patient, caregiver or mixed voice (forum 1,016; services 5,018). Following the plan, the 11 mixed posts (forum 3; services 8) are excluded from all analyses in this paper, giving 6,023 analysed kidney posts (forum 1,013; services 5,010), and counted as family-or-friend posts in a sensitivity analysis. The descriptor read covered all 6,023.

#### Posts shared with the companion study

The companion measurement study read 10,130 units with its original full-text instrument. Of the 36,564 distinct posts in this study’s frames, 338 (0.9%) were also among those units (kidney frame 198; lung cancer 38; colorectal cancer 22; ILD 1; vertigo 34; PCOS 18; obesity 39; 350 frame memberships, as some posts belong to more than one frame). They were read here under different rubrics, and no label or result of the companion study is used in this paper. Because the frames were built separately for each condition, some posts were read under more than one condition-specific rubric (1,066 reads); each read is analysed only within its own condition.

### Supplementary Methods S3. Instrument history and frozen prompts

#### Kidney rubric revision

The kidney frame was first read under the full-text rubric of the companion study (same model; here 100 posts per call, full unit text). A check by two physicians (200 posts) showed that this read rejected posts the physicians counted. The relevance rubric was rewritten with explicit inclusion and exclusion rules, the deletion test, a ‘mixed’ voice code and a recorded reason for every rejection, frozen and hashed, and every post was re-read at 10 posts per call with replies, quoted text and signatures removed. Original and revised reads agreed on confirmed status at κ 0.84 (forum 0.85; services 0.83): 581 posts were newly confirmed and 810 no longer confirmed. In a factorial experiment on 2,000 seeded kidney-frame posts crossing rubric (original, revised) with batch size (10, 100), batch size changed few labels (κ 0.94) and the rubric accounted for most changes.

#### Prompts

The relevance prompt for the six further conditions is a template whose placeholders were filled from the condition-specific definitions (Supplementary Methods S2); the full filled prompts are hashed in Table S2 (Supplementary Methods S1). Prompts are reproduced verbatim below.

#### Relevance, author and stage template (six conditions)

You are reading a public Taiwanese online post (Traditional Chinese; a forum article, or the question part of an ask-a-physician submission; any reply, answer or quoted text from another author has been removed). Read the FULL text, including the title. Do not use the name of the board or site. Answer the questions below independently of one another: decide cond_relevant without regard to who wrote the post, and decide voice without regard to whether {NAME_SHORT} is involved.

cond_relevant = 1 if {NAME_EN} ({NAME_ZH}) of the author, or of a specific person the author cares for, is a subject of the post, not merely a word in passing. It is a subject of the post if deleting every sentence that refers to it would change what the author is asking or telling; if the post would be the same post without those sentences, it is a passing mention. Count:

{COUNT}

Do not count: {EXCLUDE}; a passing mention; general information, news, advertising, sponsored content or policy that concerns no specific person; animals.

If unsure, ask: is the author asking or telling about this specific person’s {NAME_SHORT}? If yes, cond_relevant = 1.

voice = who is writing about whose health. ’patient’: the author’s own health. ’caregiver’: the health of a spouse, parent, child, other relative, partner or friend whom the author is helping, including asking a question on that person’s behalf or retelling that person’s case. ’mixed’: the author’s own {NAME_SHORT} AND a specific relative’s {NAME_SHORT} are both subjects of the post (use only when both pass the subject test; otherwise choose ’patient’ or ’caregiver’ for the person whose problem the main question is about). ’hcp’: a clinician, nurse, care worker or other professional writing about a patient in their professional care, or answering or advising others. ’media’: news, promotional, institutional, or a reposted article written by someone else. ’other’: general chat or opinion, or a second-hand story about a stranger. A question submitted to an ask-a-physician service is ’patient’ or ’caregiver’ unless its text clearly says otherwise.

reason = ’none’ if cond_relevant = 1 and voice is ’patient’, ’caregiver’ or ’mixed’; otherwise the main reason: ’passing_mention’, ’not_condition’, ’excluded_condition’, ’no_specific_person’, ’clinician_or_media_voice’, ’other’.

cond_stage describes the condition of the person whose problem is the main subject. Stage is decided only from the stated diagnosis, disease extent and prescribed treatment. Do not use whether the person is in hospital, bedridden, frail, dependent on others, or being written about by someone else. Apply the first that fits: ’advanced’ ({ADVANCED}); ’diagnosed’ ({DIAGNOSED}); ’concern’ ({CONCERN}); ’none’ (cond_relevant = 0).

quotes_value = 1 only if the author’s text states an actual number for this person relating to the condition ({VALUES}). A reference range without the person’s value, a vague description without a number, or values of unrelated tests, dates, drug doses or costs are 0. A value quoted from the care-recipient’s report counts.

#### Descriptor read (seven conditions)

Every post below has already been confirmed as a patient, family member, partner or friend writing about a specific person’s health problem. For each post, describe the person whose health problem is the main subject (for a post about both the author and a relative, the person the main question is about). Use only what the text states or clearly implies; when the text does not tell you, answer ’unclear’. Do not infer dependence or age from the fact that someone else is writing, or from the relation; use only stated facts.

subject_relation = the person’s relation to the author: ’self’, ’spouse_partner’, ’parent’ (including parent-in-law), ’child’ (including child-in-law), ’grandparent’, ’other_relative’ (sibling, grandchild, aunt, uncle, cousin), ’friend_other’, ’unclear’.

subject_age = ’child’ (under 18, or described as a baby, child, pupil or teenager), ’adult’ (18 to 64, or described as a university student, working, or young), ’older’ (65 or over, or described as elderly or as retired and over 65), ’unclear’. Do not infer age from the relation alone (a parent or grandparent may be of any age).

dependence = 1 if the text states that the person needs another person for daily activities, mobility, eating, communication or medical decisions beyond what is usual for their age, is bedridden, lives in a care home, has dementia or other cognitive impairment, or cannot speak for themselves; 0 otherwise. A healthy child is 0 (age is captured separately).

proxy_reason = ’self’ when the author is the person whose health is the subject; otherwise ’unable’ (the person is too ill, too old, cognitively impaired, a child, or cannot read or use the internet), ’delegated’ (the person could seek information but asked the author to, or the author habitually manages the family’s health matters), or ’unstated’ (no reason given).

#### Revised kidney relevance, author and context rubric

You are reading a public Taiwanese online post (Traditional Chinese; a forum article, or the question part of an ask-a-physician submission; any reply, answer or quoted text from another author has been removed). Read the FULL text, including the title. Do not use the name of the board or site. Answer the questions below independently of one another: decide ckd_relevant without regard to who wrote the post, and decide voice without regard to whether kidneys are involved.

ckd_relevant = 1 if a kidney problem of the author, or of a specific person the author cares for, is a subject of the post, not merely a word in passing. A kidney problem is a subject of the post if deleting every sentence that refers to kidneys would change what the author is asking or telling; if the post would be the same post without those sentences, it is a passing mention. Count:

(a) kidney disease of any stage, reduced kidney function, protein or blood in the urine under evaluation, dialysis (on it, starting it, or being discussed for this person), kidney transplantation;
(b) an abnormal or borderline kidney test (eGFR, creatinine, BUN, urine protein, albumin-creatinine ratio, kidney ultrasound) that the author asks or worries about;
(c) a question whether a medicine, supplement, food or symptom is harming, or signals harm to, this person’s kidneys;
(d) kidney involvement in another illness (diabetes, hypertension, lupus, gout, heart failure, cancer, including kidney cancer or removal of a kidney) when this person’s kidney function, kidney damage, dialysis or transplantation is part of what is asked or described. Kidney cancer, kidney stones, cysts, infection or blood in the urine with no mention of kidney function, kidney damage, dialysis or transplantation do not count.

Do not count: a passing mention; 腎虧/腎虛 in the traditional sense of vitality or sexual function without a kidney disease or kidney test; general information, news, advertising or policy that concerns no specific person; animals. If unsure, ask: is the author asking or telling about this person’s kidney function, kidney disease, dialysis or transplantation? If yes, ckd_relevant = 1. If the only kidney matter is a stone, an infection, cancer surgery, or sexual or general vitality (腎虧/腎虛), ckd_relevant = 0.

voice = who is writing about whose health. ’patient’: the author’s own health. ’caregiver’: the health of a spouse, parent, child, other relative, partner or friend whom the author is helping, including asking a question on that person’s behalf or retelling that person’s case. ’mixed’: the author’s own kidney problem AND a specific relative’s kidney problem are both subjects of the post (use only when both pass the subject test; otherwise choose ’patient’ or ’caregiver’ for the person whose problem the main question is about). ’hcp’: a clinician, nurse, care worker or other professional writing about a patient in their professional care, or answering or advising others. ’media’: news, promotional, institutional, or a reposted article written by someone else. ’other’: general chat or opinion, or a second-hand story about a stranger. A question submitted to an ask-a-physician service is ’patient’ or ’caregiver’ unless its text clearly says otherwise.

reason = ’none’ if ckd_relevant = 1 and voice is ’patient’, ’caregiver’ or ’mixed’; otherwise the main reason: ’passing_mention’, ’not_kidney’, ’urology_or_kidney_cancer_only’, ’shenxu_only’, ’no_specific_person’, ’clinician_or_media_voice’, ’other’.

kidney_context describes the kidney problem that is the main subject (for ’mixed’, the person the main question is about). Apply the first that fits: ’transplant’ (transplanted or being evaluated for transplantation); ’dialysis’ (on dialysis or about to start); ’diagnosed_ckd’ (established chronic kidney disease, a stated stage, or long-standing reduced function); ’aki_other’ (acute kidney injury, nephritis, a single kidney, kidney cancer or another kidney condition with function or damage at issue, without an established chronic diagnosis); ’abnormal_test’ (one or a few recent abnormal kidney results without an established diagnosis); ’concern_only’ (ckd_relevant = 1 but no kidney condition of this person is stated, for example whether a drug or supplement harms the kidneys); ’none’ (ckd_relevant = 0).

quotes_lab_value = 1 only if the author’s text states an actual number for eGFR, creatinine, BUN, urine protein, urine albumin, ACR or UPCR, or an explicit CKD stage (’stage 3’, ’第三期’, ’末期’). Values of other tests alone (uric acid, glucose,

HbA1c, potassium, phosphorus, blood pressure), a reference range without the person’s value, or ’腎功能不好’ are 0. A value quoted from the care-recipient’s report counts.

#### Kidney construct read

Full prompt as sent (base construct rubric with two added fields; Table S2). The added fields function_mentioned and decline_worry are not analysed in this paper; quotes a laboratory value comes from the kidney relevance read.

You will receive several independent posts as a JSON array of objects {"id":…, "text":…}. Apply the rubric below to EVERY post, judging each post strictly on its own text; one post must never influence another.

Every post below has already been confirmed as patient or caregiver voice. RUBRIC (constructs):

You are labelling a public Taiwanese health-forum post (Traditional Chinese) about chronic kidney disease for twelve binary constructs. Read the FULL text. Label 1 only when the construct is genuinely present for the author (for caregiver_burden, the author is a caregiver voicing burden); passing vocabulary mentions do not count. Reply ONLY with JSON exactly like:

{"info_gap":0,"cost_burden":0,"access_barrier":0,"system_nav":0,"emotional":0,"medication_exp":0,"lifestyle_burden":0, "caregiver_burden":0,"dialysis_concern":0,"transplant_concern":0,"renal_diet_burden":0,"progression_worry":0}

Core definitions (identical to the reference study): info_gap = seeks information/advice, asks a clinical question, or expresses diagnostic/treatment uncertainty. cost_burden = voices actual financial strain from care costs. access_barrier = concrete difficulty reaching care (appointments, referrals, distance, waiting). system_nav = actively working through institutional processes (insurance coverage, eligibility, paperwork, which hospital/specialty). emotional = illness-tied emotional distress (fear, despair, exhaustion). medication_exp = first-hand medication experience, efficacy doubt or side-effects. lifestyle_burden = burden from illness-driven restriction of diet, activity, work, or sleep. caregiver_burden = the author cares for a patient and voices that burden.

CKD-specific definitions: dialysis_concern = the author fears, anticipates, undergoes, or bears the burden of dialysis (haemo-or peritoneal), including vascular-access issues. transplant_concern = kidney transplantation is a real concern for the author (waiting, evaluation, donation, post-transplant). renal_diet_burden = burden or confusion specifically from renal dietary restriction (low-protein, potassium, phosphorus, fluid limits). progression_worry = worry about kidney-function decline or disease stage (eGFR, creatinine, ’第幾期’, deterioration toward dialysis). Additional fields: function_mentioned = 1 if the post mentions this person’s kidney function, a kidney test result or a CKD stage. decline_worry = 1 if the author expresses worry that this person’s kidney function will decline, progress or lead to dialysis.

OUTPUT FORMAT: for each post, output exactly one line containing one JSON object with the post’s id copied verbatim and every field, like

{"id":" <ID>","info_gap":0,"cost_burden":0,"access_barrier":0,"system_nav":0,"emotional":0,"medication_exp":0,"lifestyle_ burden":0,"caregiver_burden":0,"dialysis_concern":0,"transplant_concern":0,"renal_diet_burden":0,"progression_worr y":0,"function_mentioned":0,"decline_worry":0}

Output exactly one line per post, in the same order as the input, and nothing else (no prose, no code fences, no numbering).

#### General construct read (six conditions)

You are labelling a public Taiwanese health-forum post (Traditional Chinese) for eight binary friction constructs. Read the FULL text. Label 1 only when the construct is genuinely present for the author (or, for caregiver_burden, that the author is a caregiver voicing burden); passing mentions of vocabulary do not count. Reply with ONLY a JSON object exactly like:

{"info_gap":0,"cost_burden":0,"access_barrier":0,"system_nav":0,"emotional":0,"medication_exp":0,"lifestyle_burden":0, "caregiver_burden":0}

Definitions: info_gap = seeks information/advice, asks a clinical question, or expresses diagnostic/treatment uncertainty. cost_burden = voices actual financial strain from care costs. access_barrier = concrete difficulty reaching care (appointments, referrals, distance, waiting). system_nav = actively working through institutional processes (insurance coverage, eligibility, paperwork, which hospital/specialty). emotional = illness-tied emotional distress (fear, despair, exhaustion). medication_exp = first-hand medication experience, efficacy doubt or side-effects. lifestyle_burden = burden from illness-driven restriction of diet, activity, work, or sleep. caregiver_burden = the author cares for a patient and voices that burden.

POST:

### Supplementary Results S1. Trends in family or friend posts and the patient’s age group

**Table S5.**
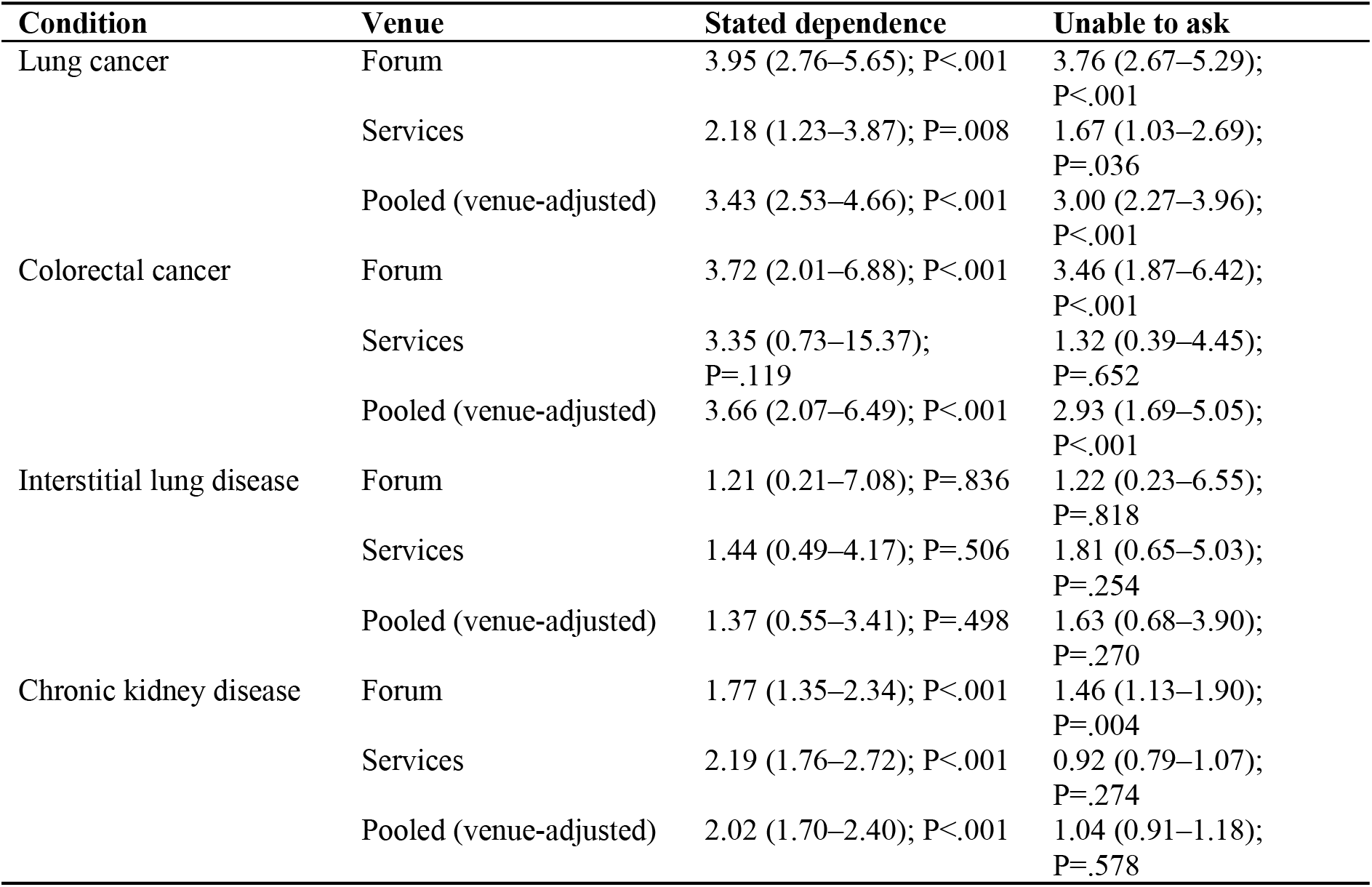
Within family or friend posts: trends across context groups (odds ratio per context group, 95% CI)

| **Condition** | **Venue** | **Stated dependence** | **Unable to ask** |
|---|---|---|---|
| Lung cancer | Forum | 3.95 (2.76–5.65); P<.001 | 3.76 (2.67–5.29); P<.001 |
|  | Services | 2.18 (1.23–3.87); P=.008 | 1.67 (1.03–2.69); P=.036 |
|  | Pooled (venue-adjusted) | 3.43 (2.53–4.66); P<.001 | 3.00 (2.27–3.96); P<.001 |
| Colorectal cancer | Forum | 3.72 (2.01–6.88); P<.001 | 3.46 (1.87–6.42); P<.001 |
|  | Services | 3.35 (0.73–15.37); P=.119 | 1.32 (0.39–4.45); P=.652 |
|  | Pooled (venue-adjusted) | 3.66 (2.07–6.49); P<.001 | 2.93 (1.69–5.05); P<.001 |
| Interstitial lung disease | Forum | 1.21 (0.21–7.08); P=.836 | 1.22 (0.23–6.55); P=.818 |
|  | Services | 1.44 (0.49–4.17); P=.506 | 1.81 (0.65–5.03); P=.254 |
|  | Pooled (venue-adjusted) | 1.37 (0.55–3.41); P=.498 | 1.63 (0.68–3.90); P=.270 |
| Chronic kidney disease | Forum | 1.77 (1.35–2.34); P<.001 | 1.46 (1.13–1.90); P=.004 |
|  | Services | 2.19 (1.76–2.72); P<.001 | 0.92 (0.79–1.07); P=.274 |
|  | Pooled (venue-adjusted) | 2.02 (1.70–2.40); P<.001 | 1.04 (0.91–1.18); P=.578 |

The ‘unable to ask’ trend was the pre-specified test (H3(ii)); stated dependence is descriptive. Stated dependence in patient-authored posts, % (95% CI): Lung cancer 1.5 (0.8–3.2); Colorectal cancer 0.1 (0.0–0.8); Interstitial lung disease 1.3 (0.2–7.0); Chronic kidney disease 1.3 (1.0–1.7); Vertigo 5.1 (4.0–6.4); Polycystic ovary syndrome 0.2 (0.1–0.5); Obesity 0.9 (0.4–1.8).

**Table S6.** Patient’s age group by author, %.

| Condition | Venue | Author | n | Child | Adult | Older adult | Unclear |
|---|---|---|---|---|---|---|---|
| Chronic kidney disease | Forum | Patient | 473 | 0.2 | 40.4 | 0.0 | 59.4 |
|  |  | Family or friend | 540 | 3.1 | 13.3 | 15.6 | 68.0 |
|  | Services | Patient | 3,522 | 1.0 | 16.5 | 0.5 | 82.1 |
|  |  | Family or friend | 1,488 | 14.2 | 17.3 | 21.2 | 47.2 |
| Vertigo | Forum | Patient | 209 | 0.0 | 32.1 | 0.0 | 67.9 |
|  |  | Family or friend | 42 | 0.0 | 19.0 | 9.5 | 71.4 |
|  | Services | Patient | 1,048 | 3.0 | 17.7 | 0.3 | 79.1 |
|  |  | Family or friend | 153 | 10.5 | 35.3 | 16.3 | 37.9 |
| Obesity | Forum | Patient | 419 | 0.5 | 63.2 | 0.0 | 36.3 |
|  |  | Family or friend | 21 | 14.3 | 42.9 | 0.0 | 42.9 |
|  | Services | Patient | 400 | 4.3 | 17.0 | 0.0 | 78.8 |
|  |  | Family or friend | 15 | 26.7 | 6.7 | 13.3 | 53.3 |
| Polycystic ovary syndrome | Forum | Patient | 217 | 0.0 | 50.7 | 0.0 | 49.3 |
|  |  | Family or friend | 11 | 0.0 | 27.3 | 0.0 | 72.7 |
|  | Services | Patient | 1,920 | 0.6 | 16.0 | 0.0 | 83.4 |
|  |  | Family or friend | 98 | 2.0 | 19.4 | 0.0 | 78.6 |
| Lung cancer | Forum | Patient | 137 | 0.0 | 45.3 | 0.0 | 54.7 |
|  |  | Family or friend | 824 | 0.0 | 10.8 | 9.6 | 79.6 |
|  | Services | Patient | 315 | 1.0 | 17.8 | 1.9 | 79.4 |
|  |  | Family or friend | 295 | 0.3 | 15.6 | 26.8 | 57.3 |
| Colorectal cancer | Forum | Patient | 134 | 0.0 | 48.5 | 0.0 | 51.5 |
|  |  | Family or friend | 327 | 0.0 | 10.7 | 7.0 | 82.3 |
|  | Services | Patient | 555 | 3.8 | 17.3 | 0.2 | 78.7 |
|  |  | Family or friend | 100 | 1.0 | 21.0 | 19.0 | 59.0 |
| Interstitial lung disease | Forum | Patient | 12 | 0.0 | 41.7 | 0.0 | 58.3 |
|  |  | Family or friend | 26 | 0.0 | 7.7 | 19.2 | 73.1 |
|  | Services | Patient | 65 | 0.0 | 23.1 | 1.5 | 75.4 |
|  |  | Family or friend | 71 | 0.0 | 14.1 | 32.4 | 53.5 |

### Supplementary Results S2. Sensitivity analyses and physician labels

**Table S7.** H1 sensitivity analyses: advanced minus concern difference, points (95% CI)

| **Condition** | **Venue** | **Primary** | **Second vendor’s own labels** | **Both readers agree on relevance, author and context group** | **Mixed counted as family** | **Age-adjusted odds ratio per context group** |
|---|---|---|---|---|---|---|
| Lung cancer | Forum | +42.1 (35.0 to 49.1); n=961 | +41.4 (30.3 to 52.6); n=379 | +42.8 (31.3 to 54.2); n=343 | +42.1 (35.0 to 49.1); n=962 | 4.13 (2.67–6.38) |
|  | Services | +61.3 (53.2 to 67.7); n=610 | +63.4 (50.3 to 72.1); n=219 | +66.7 (53.3 to 75.0); n=198 | +61.1 (52.9 to 67.5); n=611 | 4.22 (2.81–6.32) |
| Colorectal cancer | Forum | +72.1 (63.5 to 78.5); n=461 | +69.0 (54.3 to 79.4); n=168 | +68.8 (52.8 to 80.1); n=150 | +71.1 (62.5 to 77.6); n=463 | 10.08 (4.53–22.43) |
|  | Services | +65.6 (50.8 to 77.1); n=655 | +72.8 (58.7 to 82.5); n=302 | +68.5 (51.9 to 80.6); n=250 | +64.0 (49.2 to 75.6); n=664 | 9.24 (6.06–14.09) |
| Interstitial lung disease | Forum | +69.0 (38.7 to 85.2); n=38 | +52.5 (8.4 to 77.7); n=18 | +70.8 (18.2 to 87.9); n=14 | +69.0 (38.7 to 85.2); n=38 | Not estimable |
|  | Services | +54.6 (38.4 to 66.4); n=136 | +50.6 (21.2 to 69.5); n=45 | +55.0 (22.9 to 73.4); n=39 | +53.1 (36.9 to 65.1); n=137 | 8.14 (2.47–26.79) |
| Chronic kidney disease | Forum | +49.2 (42.3 to 55.4); n=1,013 | +50.9 (43.9 to 57.0); n=899 | +52.6 (45.1 to 59.1); n=783 | +49.5 (42.6 to 55.6); n=1,016 | 3.70 (2.81–4.87) |
|  | Services | +48.6 (44.8 to 52.3); n=5,010 | +53.3 (45.6 to 60.1); n=1,238 | +53.1 (44.8 to 60.5); n=1,087 | +48.6 (44.7 to 52.2); n=5,018 | 3.72 (3.06–4.53) |

**Table S8.** Kidney physician labels (200 posts; two physician authors, blinded to model labels)

| **Field** | **Model vs physician A** | **Model vs physician B** | **Physician A vs B** |  |
|---|---|---|---|---|
| Kidney post (relevance) | 0.18 | 0.38 | 0.36 |  |
| Family or friend authorship | 0.73 | 0.76 | 0.93 |  |
| Context group | 0.48 | 0.58 | 0.69 |  |
| **Labels (139 posts from strata not selected on author or context)** | **Concern, % (95% CI)** | **Diagnosed** | **Advanced** | **Trend P** |
| Physician A’s own labels | 24 (13–41); n=33 | 22 (12–38); n=36 | 47 (33–61); n=47 | .023 |
| Physician B’s own labels | 14 (6–31); n=28 | 33 (20–48); n=40 | 48 (33–63); n=40 | .004 |
| Model labels on the same posts | 13 (5–29); n=31 | 43 (28–59); n=35 | 71 (51–85); n=24 | — |

The physicians labelled under a broader worksheet definition of a kidney post (kidney disease, kidney test result, dialysis or transplant). Of 145 posts the model confirmed, 142 were counted by at least one physician. Re-reading the 200 posts under the physicians’ own worksheet definition (both vendors) changed agreement on relevance little (κ gpt-6.1-sol: 0.21 and 0.44, Claude Sonnet 5.5: 0.23 and 0.40), indicating disagreement about which posts are about kidney disease, not only a definitional difference. These labels prompted the rubric revision (in aggregate) and are therefore not an independent check of it.

**Table S9.**
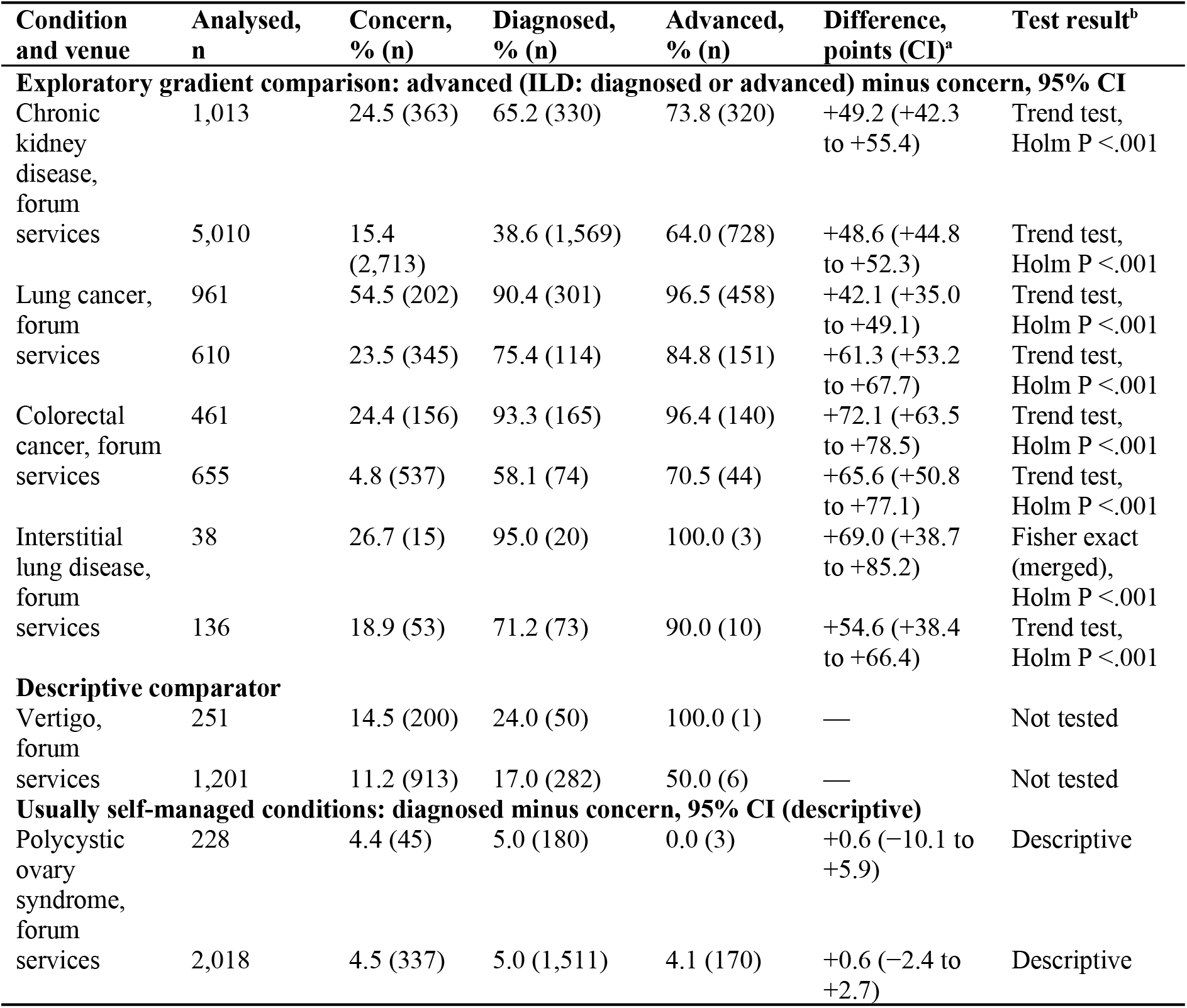
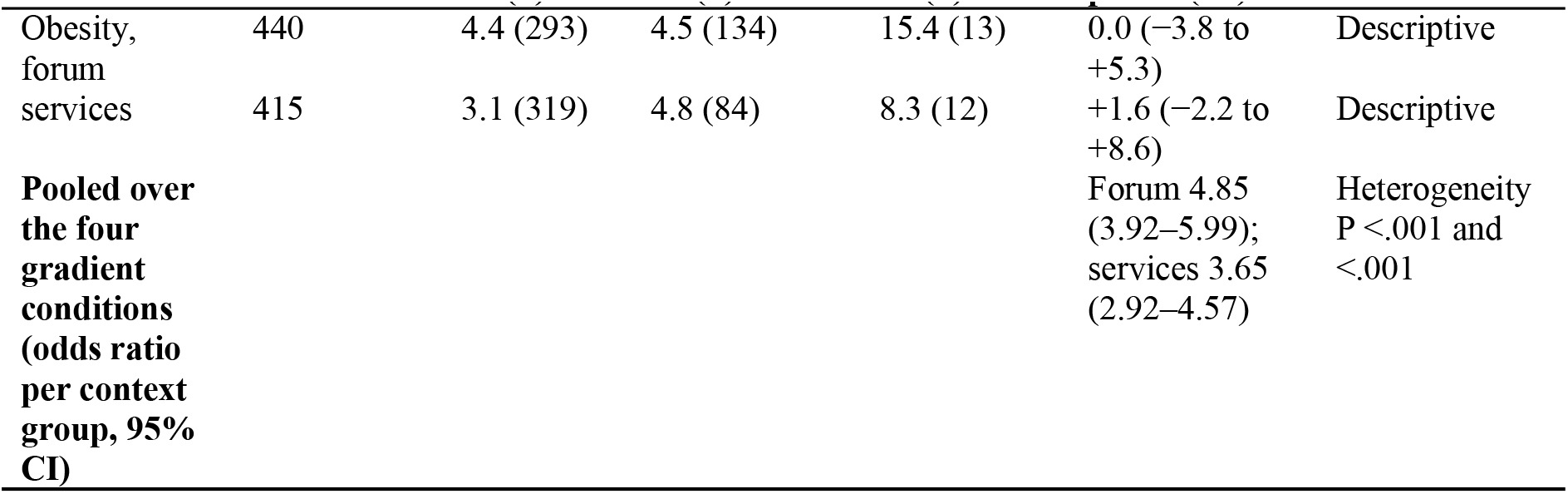
Family authorship by context group in kidney disease and six comparison conditions (exploratory; comparisons specified after the pattern had been seen)

| Condition and venue | Analysed, n | Concern, % (n) | Diagnosed, % (n) | Advanced, % (n) | Difference, points (CI)<sup>a</sup> | Test result<sup>b</sup> |
|---|---|---|---|---|---|---|
| **Exploratory gradient comparison: advanced (ILD: diagnosed or advanced) minus concern, 95% CI** |  |  |  |  |  |  |
| Chronic kidney disease, forum | 1,013 | 24.5 (363) | 65.2 (330) | 73.8 (320) | +49.2 (+42.3 to +55.4) | Trend test, Holm P <.001 |
| services | 5,010 | 15.4 (2,713) | 38.6 (1,569) | 64.0 (728) | +48.6 (+44.8 to +52.3) | Trend test, Holm P <.001 |
| Lung cancer, forum | 961 | 54.5 (202) | 90.4 (301) | 96.5 (458) | +42.1 (+35.0 to +49.1) | Trend test, Holm P <.001 |
| services | 610 | 23.5 (345) | 75.4 (114) | 84.8 (151) | +61.3 (+53.2 to +67.7) | Trend test, Holm P <.001 |
| Colorectal cancer, forum | 461 | 24.4 (156) | 93.3 (165) | 96.4 (140) | +72.1 (+63.5 to +78.5) | Trend test, Holm P <.001 |
| services | 655 | 4.8 (537) | 58.1 (74) | 70.5 (44) | +65.6 (+50.8 to +77.1) | Trend test, Holm P <.001 |
| Interstitial lung disease, forum | 38 | 26.7 (15) | 95.0 (20) | 100.0 (3) | +69.0 (+38.7 to +85.2) | Fisher exact (merged), Holm P <.001 |
| services | 136 | 18.9 (53) | 71.2 (73) | 90.0 (10) | +54.6 (+38.4 to +66.4) | Trend test, Holm P <.001 |
| **Descriptive comparator** |  |  |  |  |  |  |
| Vertigo, forum | 251 | 14.5 (200) | 24.0 (50) | 100.0 (1) | — | Not tested |
| services | 1,201 | 11.2 (913) | 17.0 (282) | 50.0 (6) | — | Not tested |
| **Usually self-managed conditions: diagnosed minus concern, 95% CI (descriptive)** |  |  |  |  |  |  |
| Polycystic ovary syndrome, forum | 228 | 4.4 (45) | 5.0 (180) | 0.0 (3) | +0.6 (−10.1 to +5.9) | Descriptive |
| services | 2,018 | 4.5 (337) | 5.0 (1,511) | 4.1 (170) | +0.6 (−2.4 to +2.7) | Descriptive |
| Obesity, forum | 440 | 4.4 (293) | 4.5 (134) | 15.4 (13) | 0.0 (−3.8 to +5.3) | Descriptive |
| services | 415 | 3.1 (319) | 4.8 (84) | 8.3 (12) | +1.6 (−2.2 to +8.6) | Descriptive |
| **Pooled over the four gradient conditions (odds ratio per context group, 95% CI)** |  |  |  |  | Forum 4.85 (3.92–5.99); services 3.65 (2.92–4.57) | Heterogeneity P <.001 and <.001 |

Shares are percentages of analysed posts (n per context group in parentheses; model estimates); advanced cells with fewer than 20 posts were merged with the diagnosed group for the ILD trend tests, and those in PCOS (forum), obesity and vertigo were not tested or interpreted; the forum ILD concern cell (15 posts) was retained as the reference. Context-group definitions are condition-specific (Supplementary Methods S2). ^a^Gradient comparison: advanced (ILD: diagnosed or advanced, merged) minus concern, Newcombe 95% CI. Self-managed conditions: diagnosed minus concern, Newcombe 95% CI, shown descriptively; for PCOS on the services, advanced minus diagnosed was −0.9 (95% CI −3.3 to 3.3). ^b^Risk differences compare category endpoints, whereas the trend test assesses the fitted ordinal trend (groups scored 0, 1, 2), Holm-adjusted across the 8 cells; forum ILD: Fisher exact test of the merged contrast, because the adjusted logistic model showed complete separation. ILD, interstitial lung disease; PCOS, polycystic ovary syndrome.

**Supplementary Figure S1.**
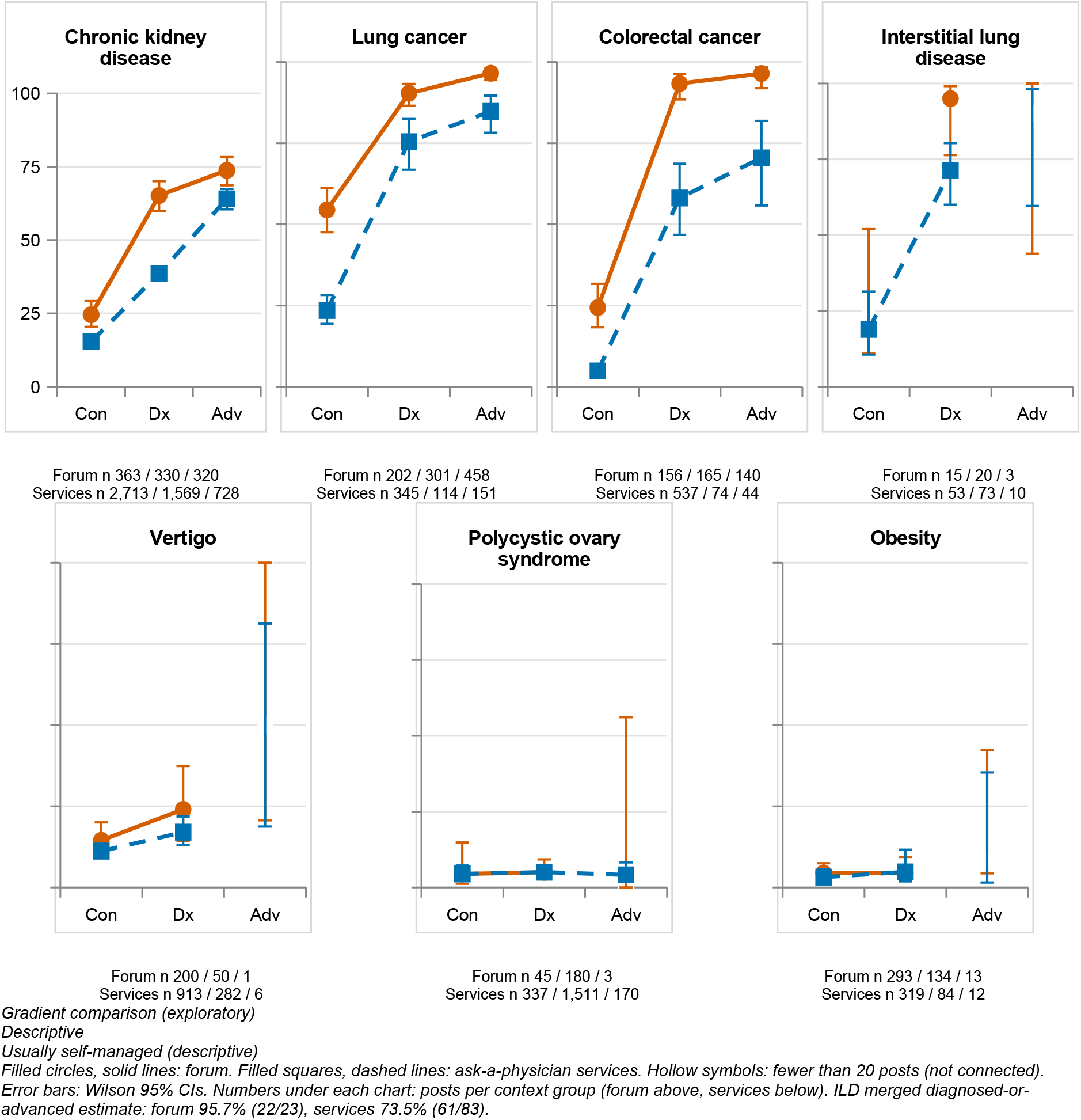
Family or friend authorship by context group in kidney disease and six comparison conditions.

Forum (circles, solid lines) and ask-a-physician services (squares, dashed lines), with Wilson 95% CIs, at the concern (Con), diagnosed (Dx) and advanced (Adv) context groups (model labels; cross-sectional). Hollow symbols mark cells with fewer than 20 posts, which are not connected and not interpreted, except the forum interstitial lung disease concern cell (15 posts), retained as the reference cell of its test; diamonds in that panel show the merged diagnosed-or-advanced estimate. Numbers beneath each panel are posts per group (forum above, services below). Conditions are grouped as in the analysis plan, whose comparisons are exploratory because the pattern had been inspected before the plan was frozen.

### Supplementary Checklists. STROBE and TRIPOD-LLM

**Table S10.** STROBE checklist (cross-sectional studies)

| Item | Recommendation | Location |
|---|---|---|
| 1 | Title and abstract: design indicated; informative, balanced summary | Title; Abstract |
| 2 | Background and rationale | Introduction |
| 3 | Objectives, including prespecified hypotheses | Introduction (final paragraph); Methods, Design and Timeline |
| 4 | Study design | Methods, Design and Timeline |
| 5 | Setting, locations, dates | Abstract (posts 1995–2026); Methods, Kidney Frame and Reading (reads October 2026) |
| 6 | Eligibility and selection | Methods, Kidney Frame and Reading; Supplementary Methods S2 |
| 7 | Variables: outcomes, exposures, effect modifiers | Methods, Friction Framework and Kidney Fields; Kidney Frame and Reading (author, context group); Outcomes and Statistical Analysis |
| 8 | Data sources and measurement | Methods, Kidney Frame and Reading; Replication and Physician Labels; Supplementary Methods S3 |
| 9 | Bias | Methods (frozen rubrics; second vendor; physician labels; timeline); Discussion, Limitations |
| 10 | Study size | Methods, Kidney Frame and Reading (full read); Outcomes and Statistical Analysis (comparison conditions, seeded samples) |
| 11 | Quantitative variables | Table 4 footnote (context group scored 0, 1, 2); Table S3 and Table S9 (merging rule) |
| 12 | Statistical methods, subgroups, missing data, sensitivity analyses | Methods, Outcomes and Statistical Analysis; Supplementary Methods S1 (deviations) |
| 13 | Numbers at each stage of the study; flow | Results, Kidney-Related Posts; Table 1; Supplementary Methods S2 (flow of posts) |
| 14 | Descriptive data | Table 1; Supplementary Results S1 |
| 15 | Outcome data | Tables 2–4; Figures 1 and 2 |
| 16 | Main results with precision; adjusted estimates | Results; Tables 2–4 |
| 17 | Other analyses | Results, Whose Needs and Burdens; Who Writes, by Clinical Context (sensitivity); Supplementary Results S2 |
| 18 | Key results | Discussion (first paragraph) |
| 19 | Limitations | Discussion, Limitations |
| 20 | Interpretation | Discussion |
| 21 | Generalisability | Discussion, Limitations (one health system and language) |
| 22 | Funding | Acknowledgments |

**Table S11.** TRIPOD-LLM items applicable to LLM-based text annotation.

| Item | Description | Location |
|---|---|---|
| LLM identification | Model names, versions, access route, dates | Methods, Kidney Frame and Reading (gpt-6.1-sol, command-line interface, October 2026); Replication and Physician Labels (Claude Sonnet 5.5) |
| Task | Relevance, author, context group, descriptor and content labelling of posts | Methods, Kidney Frame and Reading; Friction Framework and Kidney Fields |
| Prompting | Full prompts, frozen and hashed before use | Supplementary Methods S3 (verbatim); Supplementary Methods S1 (freeze times; hashes on request) |
| Prompt development | Development data and revision history | Supplementary Methods S3 (kidney rubric revision) |
| Data | Sources, frames, sampling, preprocessing | Methods, Data Sources; Kidney Frame and Reading; Supplementary Methods S2 |
| Evaluation | Second-vendor agreement; physician labels; agreement measures | Methods, Replication and Physician Labels; Table 1; Supplementary Results S2 |
| Performance | Agreement by condition and venue with uncertainty | Tables 1 and 2 (second-vendor κ by venue and field, kidney posts; no intervals); Supplementary Results S2 (sensitivity analyses) |
| Limitations | Reproducibility versus validity; training-data overlap | Discussion, Limitations |
| Availability | Prompts, labels and code | Supplementary Methods S3 (prompts); Data Sharing Statement (data on request) |

